# Evaluation of reported claims of sex-based differences in treatment effects across meta-analyses: A meta-research study

**DOI:** 10.1101/2024.07.04.24309572

**Authors:** Lum Kastrati, Sara Farina, Angelica-Valz Gris, Hamidreza Raeisi-Dehkordi, Erand Llanaj, Hugo G. Quezada-Pinedo, Lia Bally, Taulant Muka, John P.A. Ioannidis

## Abstract

**Importance:** Differences in treatment effects between men and women may be important across diverse interventions and diseases.

**Objective:** We aimed to evaluate claims of sex-based differences in treatment effects across published meta-analyses.

**Data Sources:** PubMed (searched up to January 17, 2024).

**Study Selection:** Published meta-analyses of randomized controlled trials (RCTs) that had any mention of sex (male/female) subgroup or related analysis in their abstract.

**Data Extraction and Synthesis:** We determined how many meta-analyses had made claims of sex-based differences in treatment effects. These meta-analyses were examined in depth to determine whether the claims reflected sex-treatment interactions with statistical support or fallacious claims and categorized the frequency of different fallacies. For claims with statistical support, we examined whether they were considered and discussed in UpToDate. Whenever possible, we re-analyzed the p-value for sex-treatment interaction.

**Main Outcomes and Measures:** Number of claims with statistical support and fallacious claims; clinical implications of subgroup differences.

**Results:** 216 meta-analysis articles fulfilled the eligibility criteria. Of them, 99 stated in the abstract that that there was no sex-based difference, and 20 mentioned a sex-based subgroup analysis without reporting results in the abstract. 97 meta-analyses made 115 claims of sex-based differences. Of them, 27 claims across 21 articles had statistical support at p<0.05. 4/27 claims were mentioned in UpToDate, but none led to different recommendations for men and women. 35 articles had 39 fallacious claims where the sex-treatment interaction was not statistically significant (significant effects in one sex (29 claims in 25 articles), larger effects in one sex (7 claims in 7 articles), other (3 claims in 3 articles)). Another 44 articles made claims based on potentially fallacious methods (39 based on meta-regression of percentage of one group and 5 providing the results of only one group), but proper data were unavailable to assess statistical significance.

**Conclusions and relevance:** Few meta-analyses of RCTs make claims of sex-based differences in treatment effects and most of these claims lack formal statistical support. Statistically significant and clinically actionable sex-treatment interactions may be rare.

## INTRODUCTION

Treatment effect differences between men and women are an important topic across very different interventions and diseases. Various regulatory initiatives have attempted to increase female participation in clinical studies and improve the quality and reporting of study results stratified by sex [1–3]. Exploring differences between groups that could potentially modify patient management has also received impetus with the advent of personalized medicine [4–6]. However, conducting and reporting sex-realted subgroup analyses for randomized controlled trials (RCTs) is plagued by unaccounted multiplicity in testing, not pre-specifying subgroup analyses, protocol deviations, and failure to provide statistical support for subgroup claims [7–9]. Oftentimes, a subgroup difference is claimed when the intervention is significant in only one group or when one group shows a larger point estimate without formally testing sex-treatment interaction [10, 11]. An evaluation of Cochrane reviews found that statistically significant sex-treatment interactions in RCTs were only slightly more frequent than expected by chance and there was scarce evidence of subsequent corroboration or clinical relevance of claimed sex-treatment interactions [12]. Misleading claims of sex-based subgroup differences can result in erroneous clinical decision-making [5, 13–15]. This may be even more relevant, when these claims come from systematic reviews and meta-analyses. Meta-analyses are influential for shaping guideline recommendations, but may have major flaws [16].

Here we performed an empirical meta-research study to evaluate sex-based claims of treatment effect differences reported in meta-analyses of RCTs across the medical literature. We focused on claims made in the abstracts of meta-analyses.

## METHODS

### Overall design

The original protocol for this meta-research project has been registered in OSF (https://osf.io/r4zwq/). We assembled published meta-analyses of RCTs that had any mention of sex (male/female, men/women) subgroup or related analysis in their abstract. Among these meta-analyses, we determined how many made claims of sex-based differences in treatment effects in their abstracts. These meta-analyses were further examined in-depth to determine whether these claims reflect sex-treatment interactions with statistical support at the p<0.05 level. For those that lacked such statistical support, we categorized the frequency of different fallacies that they represented. For those that had statistical support, we also searched to identify the most recent updated meta-analysis on the same topic; and whether the sex-treatment interaction was still seen in the updated meta-analysis. Finally, for sex-treatment interactions with statistical support, we examined whether they were discussed in UpToDate, a widely used evidence-based reference source [17] and whether they led to recommendations for differential treatment in men and women.

### Search strategy and eligibility criteria

We used PubMed to identify meta-analyses that have in their title and/or abstract any mention of subgroup along with any sex-based terms and are likely to represent meta-analyses of randomized trials (with or without inclusion of non-randomized data). The search string was “randomi* AND subgroup [tiab] AND ( sex [tiab] OR gender [tiab] OR (men [tiab] OR women [tiab]) OR (male [tiab] OR female [tiab])). The search was with Systematic Review and Meta-analysis filters in PubMed. Last update was done on January 17, 2024.

All retrieved results were screened to exclude studies addressing only one specific sex. Among those considering both sexes, studies were included if they were indeed systematic reviews and meta analyses of RCTs; they made any sex-based statement regarding any treatment effect in the abstract or promised in the abstract to perform subgroup analyses by sex; they pertained to humans; and were published in English language.

### Categorization of meta-analyses

Eligible meta-analyses were categorized into:

1. Studies that mentioned in the abstract a plan to perform some sex-based subgroup analysis for treatment effects, but reported no relevant results in the abstract.
2. Studies that reported that there was no difference (or results were similar) between men and women.
3. Studies that made some sex-based claim about treatment effects in the abstract.

Studies in the first two groups were only described and not pursued further. For studies that made any sex-based claim about treatment effects in the abstract (group III), we determined whether these claims are based on statistically significant (p<0.05) sex-treatment interaction upon subgroup analysis, or they represent a fallacy.

### Data extraction and further assessments for sex-treatment interactions with statistical support

For claims that had statistical support for sex-treatment interaction, we determined the following by perusing the respective meta-analysis article in full text:

1. whether the subgroup analysis was performed for the primary outcome, a secondary outcome, an outcome of unspecified priority (the meta-analysis does not clarify which outcome(s) were primary), or for multiple outcomes (and if so, whether these were primary, secondary, or unspecified) in the meta-analysis;
2. the p-value for the difference of treatment effects; if this was not already calculated by the meta-analysis authors, we calculated it. We grouped p-values into suggestive (between 0.05 and 0.005) and fully significant (p<0.005) [18].
3. whether (and how) the authors of the meta-analysis discussed the clinical significance (if any) of this finding.

We then searched in PubMed for more recent meta-analyses on the same topic. We fused the “Cited by” function to find any citation by a more recent meta-analysis. Whenever no more recent meta-analysis was found by this approach, we also searched using the “Systematic Review” and “Meta-analysis” filters and the name(s) of the treatment intervention(s) and the disease of interest. The most recently published meta-analysis was examined in full text to identify assessments of the sex-based claim including also newer data. Whenever this analysis had not been done, we performed the analysis of subgroup differences ourselves using the same model as in the index meta-analysis.

Moreover, we searched the most recent release of UpToDate [17] to see if there is any mention of the sex-treatment interaction, and, if so, whether any recommendations were made for treating differently men and women.

Finally, as a late addition to our protocol, for claims with statistical support, we checked whether these meta-analyses mentioned a published protocol (in journal or online) and, if so, whether the protocol pre-specified the subgroup analysis.

### Categorization of potentially fallacious claims

Claims of sex-based differences that were potentially fallacious were categorized into the following categories, based on what the meta-analysis authors stated in the abstract:

1. Meta-regression claims using percentage of men and women in each trial as a regression variable, or other claims based on percentage of men and women in each trial (e.g. “a benefit was seen in trials with >60% women”) – these claims are subject to ecological fallacy.
2. Nominal statistical significance (p <0.05) of the treatment effect was present on one sex group but not on the other.
3. One group had larger point estimate.
4. Treatment effects in opposite directions.
5. Other potential fallacy.

All these claims may or may not also have statistical support for sex-treatment interaction. Whenever possible, we examined whether these articles provided sufficient information to calculate a formal sex-treatment interaction p-value. Those where p<0.05 was obtained were considered under the group of those with statistical support, as above. Those with p>=0.05 were considered fallacies. Claims not providing sufficient information to calculate a p-value for sex-treatment interaction were considered in a separate category.

### Data extraction process

For all the above-mentioned steps, screening, data extraction and classification of the claims were performed by independent reviewers. Potential discrepancies were arbitrated by a third reviewer (JPAI).

We extracted the reported evidence synthesis method used to explore subgroup differences and the results. For those meta-analyses that claimed a difference but did not perform a formal subgroup analysis, we extracted the data, when available, and calculated the p-value for sex-treatment interaction.

The following additional characteristics were extracted by one data extractor (LK) and reviewed by two additional independent reviewers (SF, AVG): Author, condition/disease, compared treatment interventions, primary outcome, outcome of the claimed difference, method used to claim difference, number of events and participants for intervention and control for both groups. When multiple outcomes on the same comparison and disease were eligible and provided data to evaluate sex-based differences, priority was given to the primary outcome. When there were multiple primary outcomes with sex-based differences, we captured all of them.

### Statistical analysis

For consistency, studies with metrics based on binary data were standardized to risk ratios and studies with continuous outcomes were handled as mean differences. In each meta-analysis, estimates on men from each trial were combined; the same was done for women; and then summary estimates were compared. We used the DerSimonian-Laird random effects model [19]. As sensitivity analysis, we also used fixed effects analysis. Statistically significant sex-treatment interactions were also retested with Hartung-Knapp-Sidik-Jonkman (HKSJ) random effects. HKSJ often outperforms the DerSimonian-Laird method [20]. When only pooled estimates for men and for women were available in the meta-analysis articles, we used these reported estimates to derive the p-value for sex-treatment interaction [21]. Analyses were run in STATA.

## RESULTS

### Eligible meta-analyses

After removing duplicates (n=165), 1568 articles were available for screening. Eventually 216 articles were eligible and made statements about sex-based differences in the abstract (Figure 1). Of them, 99 articles stated that there was no sex-based difference, and 20 mentioned a sex-based subgroup analysis but did not report results in the abstract (Table S1&S2). The remaining 97 meta-analyses made 115 claims of the presence of sex-based differences.

**Figure 1.**
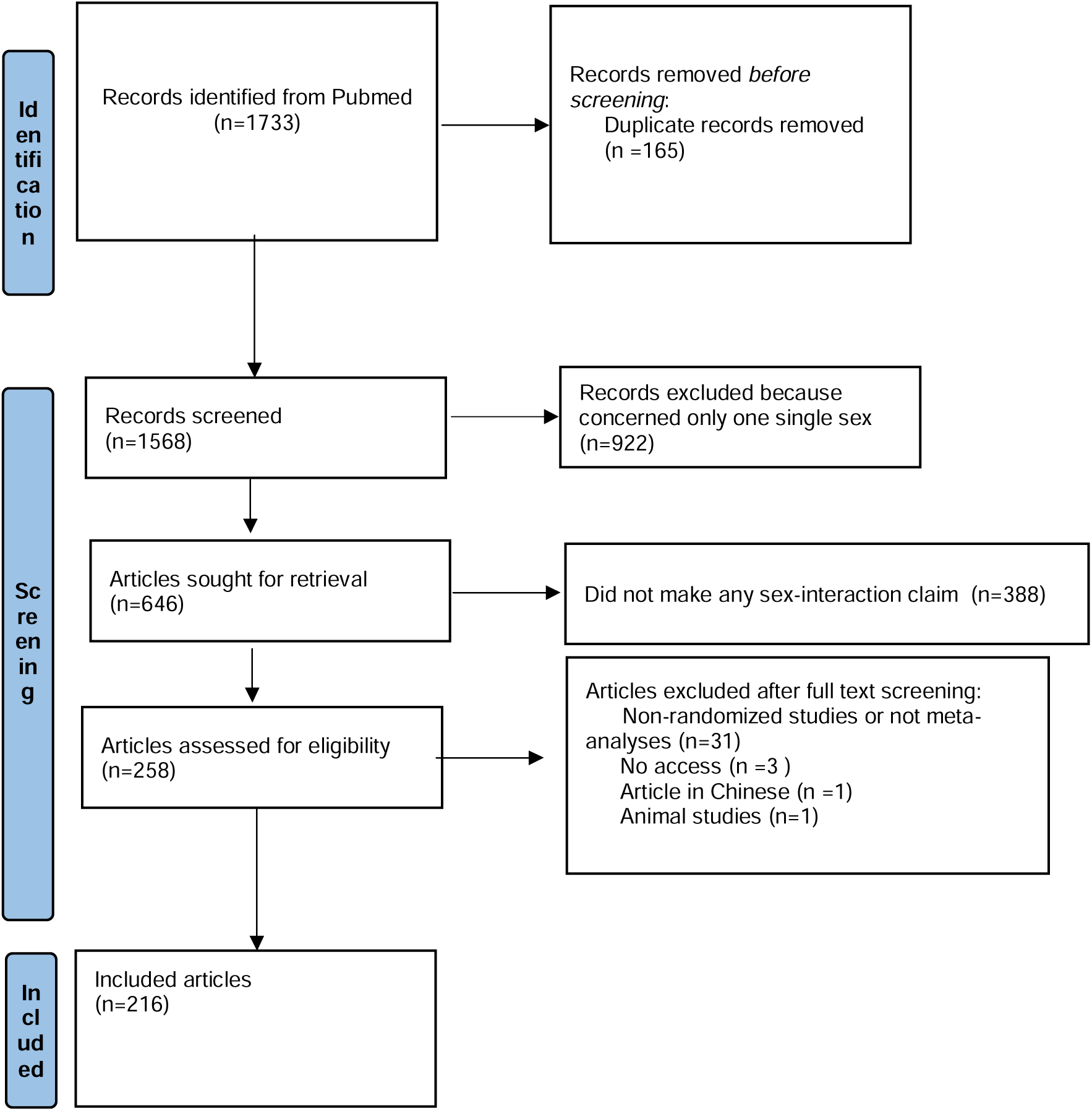
Flow-chart of the selection of the meta-analysis articles.

### Claims with statistical support

Among the 97 articles with 115 claims of sex-based differences, 21 articles had 27 claims with statistical support at p<0.05 (Table 1, Table S3) [22–42]. We had to calculate the p-value of the difference from provided pooled estimates for 12/27 claims. 11/27 claims had p<0.005.

**Table 1.** Characteristics of the 27 claims (in 21 articles) with statistical support at p<0.05.

| **Characteristics** | **No. Articles** | **No. Claims** |
| --- | --- | --- |
| Claims for the primary outcome | 16/21 | 24/27 |
| P value for sex-treatment interaction provided by authors | 11/21 | 15/27 |
| Information to calculate the p-value, when P value was not reported | 4/21 | 6/27 |
| Not enough information for exact P-value calculation but derived the p-value from the pooled estimates for each group* | 17/21 | 21/27 |
| Statistical support with a p-value for interaction $< 0.05$, but $\geq 0.005$ | 13/21 | 16/27 |
| Statistical support with a p-value for interaction $< 0.005$ | 10/21 | 11/27 |
| Authors discuss the clinical significance of the difference | 3/21 | 4/27 |
| Updated meta-analysis available | 1/21 | 1/27 |
| Mentioned in UpToDate | 3/21 | 4/27 |

\*The authors only provide the pooled estimates and respective confidence intervals

Only 4/21 articles had sufficient data allowing re-analysis (Table 2, Figure S1-S4b) [23, 35, 40, 42]. However, only one Cochrane meta-analysis had p<0.05 for all the three meta-analysis models (DerSimonian-Laird, HSJK, fixed effects) [23]; the other three meta-analyses had p>0.05 when analyzed by the HSJK model (Table 2).

**Table 2.** Re-analysis of meta-analyses where the authors had declared p<0.05 for sex-based subgroup differences.

| Meta-analysis | Outcome | Reported P-value | Reported Model | P-value after Re-analysis |  |  | Re-analysis |
| --- | --- | --- | --- | --- | --- | --- | --- |
|  |  |  |  | D-L | HKSJ | FE |  |
| Shu[35] | Adiponectin | 0.03 | RE | 0.19 | 0.18 | 0.03 | Fig. S6 |
| Zhao[40] | FPG | 0.049 | Inverse variance weighting with RE | 0.08 | 0.14 | 0.02 | Fig S7a |
|  | HOMA-IR | 0.03 | Inverse variance weighting with RE | 0.03 | 0.05 | 0.03 | Fig S7b |
| Chambers[23] | Stroke/death | 0.08 | Fixed Effects | 0.01 | 0.01 | 0.01 | Fig S8 |
| Xie [67] | MI | 0.02 | D-L | 0.03 | 0.09 [78] | 0.00 | Fig S9a |
|  | Stroke | 0.01 | MH | 0.01 | 0.12 | 0.01 | FigS9b |

**D-L-** DerSimonian and Laird
**HKSJ-** Hartung- Knapp-Sidik-Jonkman
**FE-**Fixed Effects- Mantel Hanzel for binary outcomes and Inverse Variance for Continuous
**RE-** Random Effects
**FPG-** Fasting Plasma Glucose
**HOMA-IR** – Insulin Resistance
**MI-** Miocardial Infarction
**MH-** Mantel Haenzel

Upon full-text perusal, clinical significance was discussed in three meta-analyses [23, 30, 31], while the other 18 articles reported it was hard to explain the sex differences or recommended more studies. Li et. al. [31] succinctly mentioned that the differences can lead to significantly different improved immunotherapy efficacy of PD-PD-L1 in progression-free and overall survival for women for non-small cell lung carcinoma, with their results being inconsistent with previous literature. Kazemi et. al. briefly mentioned that probiotics can help increase CD4 count specifically in women with HIV-1 infection [30]. Chambers et. al. delved deeper into explaining that women with asymptomatic carotid stenosis might not benefit from carotid enterectomy, but also mentioned that women subgroups had smaller sample sizes and shorter follow-ups, and also acknowledged the possibility for type II error [23].

Among claims with statistical support, 10/21 meta-analyses (15/27 claims) had a protocol (PROSPERO n=8, OSF n=1, manuscript supplement n=1). 6 of them had prespecified the subgroup analysis in the protocol (Table S3).

Upon searching for updated meta-analyses on the 27 claims with statistical support, only one updated meta-analysis [26] had included additional, more recent data: Purslane was still associated with significantly lower fasting blood glucose in women versus men (p<0.001) [43].

Findings from only 3 of the 27 claims were mentioned in UpToDate. However, suggestions for different management for men and women were not made for any (Table 3). Findings from [23] were acknowledged in UpToDate, but they declared no difference on managing asymptomatic carotid stenosis regarding sex [44]. UpToDate similarly only acknowledged [45] larger benefits of sigmoidoscopy for incidence and mortality of colorectal cancer for men [29]. Lastly, the significantly higher benefit of rituximab on event-free survival for diffuse large B-cell lymphoma in women versus men [41] was mentioned along with the physiological rationale, with no implications for different treatment strategies [46].

**Table 3.** Characteristics of the claims of sex-based differences with statistical support that are also mentioned in UpToDate.

| Condition | Intervention | Control | Outcome | Results | Numerical effect and 95%CI |  | P-value |
| --- | --- | --- | --- | --- | --- | --- | --- |
|  |  |  |  |  | Male | Female |  |
| Colorectal cancer | Faecal blood testing | Sigmoidoscopy/ colonoscopy | Cancer incidence | Sigmoidoscopy had a greater effect in men, for colorectal cancer incidence | 0.75 (0.71-0.79)* | 0.86 (0.81-0.92)* | 0.001 |
|  |  |  | Cancer mortality | Sigmoidoscopy had a greater effect in men for mortality | 0.67 (0.61-0.75)* | 0.85(0.75-0.96)* | 0.015 |
| Asymptomatic carotid stenosis | Carotid Endarterectomy | Medication | Perioperative stroke or death or subsequent carotid stroke | Men benefited more than women from carotid endarterectomy | 0.49 (0.36, 0.66)* | 0.96 (0.64, 1.44)* | 0.008 |
| Diffuse large B-cell lymphoma | Rituximab | Observation | Event-free survival | Men have longer significantly different event-free survival than women | 0.53 (0.34, 0.82)† | 0.99( 0.64, 1.52) † | 0.048 |

Estimates denoted with \* are Risk Ratios Estimates denoted with † are Hazard Ratios

### Fallacious claims

35 articles had 39 fallacious claims (Table 4, Table S4). The most common form of fallacy (25 articles, 29 claims) was encountered in studies that stated statistically significant benefit/harm for only one sex, but the difference between the two groups was not statistically significant [26, 28, 47–69]. In 7 articles (7 claims) the effect was stated to be larger in one group, but the overall differences were not statistically significant [33, 70–75]. 3 articles (3 claims) had different forms of fallacies: a p-value of 0.09 was considered significant [76]; the authors presented a p-value of 0.002, however, after re-calculating the p-value from reported data, we obtained p=0.12 [77]; and one study presented the p-value for comparing male vs female vs mixed.

**Table 4.** Characteristics of articles with claims of sex-based differences made in a fallacious way.

| Characteristics |  | No. Articles<br>N=35 | No. Claims<br>N=39 |
| --- | --- | --- | --- |
| Fallacy category | Significant effect only in one sex without true difference | 25 | 25 |
|  | One group had larger estimates without true difference | 7 | 7 |
|  | Other forms of fallacies | 3 | 3 |
| Provided a P-value for sex-treatment interaction |  | 8 | 8 |
| Provided results per sex for each trial |  | 5 | 5 |
| Not enough information for exact P-value calculation but derived the p-value from the pooled estimates for each group* |  | 20 | 24 |

\*The authors only provide the pooled estimates and respective confidence intervals

In 8 of 35 articles (involving 8/39 claims) the authors provided also the p-value for sex-based subgroup differences (all p>0.05) [48, 50, 59, 62, 72, 76–78].

Overall, only 5 of the 35 articles (5 claims) presented results separately for men and women from each trial [50, 52, 59, 72, 78]. Upon re-analysis, these studies had p>0.05 by both random and fixed effects (Figure S5-S9). Another 30 of the articles (34 claims) provided only pooled estimates for each sex group. Upon calculating the p-value for differences between groups using the provided pooled, estimates, 3 articles had both claims with statistical support and others without statistical support [26, 28, 33]; all other studies had p≥0.05.

### Claims without the ability to test for statistical support

44 articles made 49 claims that were potentially fallacious but no data were provided to allow probing their statistical support (Table S5). In 39 articles, 44 claims were based on ecological meta-regression or similar approaches based on percentage of men or women in each trial [79–117]. In 4 other articles (4 claims), authors reported estimates of only one subgroup [118–121], and in one other case the authors based their claim solely on the majority of participants being male [122].

## DISCUSSION

This meta-research evaluation identified fewer than 100 meta-analyses of RCTs that made claims for sex-based differences of treatment effects in their abstracts. This is a very small proportion among the tens of thousands of published meta-analyses indexed in PubMed. Furthermore, most reported claims for sex-based differences for treatment effects either lack statistical support even at a very lenient p<0.05 threshold; or are made in potentially fallacious ways and no sufficient data are provided to assess their statistical support. Finally, even these claims that have statistical support do not seem to affect differentially the management of the respective conditions in men versus women.

We identified many different ways in which claims for sex-based differences are made in potentially fallacious ways. The most common pattern involved meta-regression using proportion of men or women as an independent variable or, similarly, claims based on the percentage of one of the two groups. Meta-regression is a convenient method, but presents methodological pitfalls and ecological fallacy is a major problem [123, 124]. The second most common pattern was when researchers focused on the presence of statistical significance in one sex but not the other. These are common misleading approaches in interpreting subgroup analyses in RCTs, meta-analyses thereof, and various other types of medical research [7, 13, 125–127]. They reveal a persistent problem with the literature of subgroup differences and its misleading implications.

In trying to explore subgroup differences, meta-analyses face the challenge that reported patient characteristics from the primary RCTs may be limited [128, 129]. Reporting deficiencies are further exaggerated in meta-analyses. Very few meta-analyses provided sufficient trial-level information separately for each sex to allow proper re-analysis of subgroup claims.

A prior evaluation of sex-based differences across the entire Cochrane database [12] identified only 41 reviews that contained relevant information that would allow assessing sex-treatment interaction. The same (or worse) missingness of relevant sex-specific data may apply also to journal-published meta-analyses besides Cochrane. Cochrane meta-analyses are usually more thorough and detailed than other journal-based meta-analyses [130, 131]. The only meta-analysis in our sample with p<0.05 after re-analysis with the HKSJ method, was actually a Cochrane review [23]. It is possible that several additional meta-analyses may have performed analyses of sex-based differences but did not report anything in their abstracts. However, the proportion among them that might have had statistically significant differences is probably much lower than what we saw in the examined sample. Abstracts tend to select for the most interesting and significant results, and we focused on abstract-listed claims exactly for that purpose.

Empirical epidemiological assessments of systematic reviews and meta-analyses suggest that about half perform some subgroup analyses and about one in ten performs also meta-regressions [132]. While sex is an important variable to consider, sex-based subgroup analyses are not easy to perform given the lack of reporting of sufficient information per trial. Nevertheless, even in meta-analyses of individual-level data where subgroup analyses are almost always performed [133] statistically significant sex-based differences are uncommon. Across 327 meta-analyses of individual-level data from RCTs, only 8 found statistically significant sex-treatment interactions [133] It should be acknowledged that even meta-analyses may have limited power to detect sex-based differences [134]. However, the emerging evidence is compatible with the possibility that sex-treatment interactions are indeed very uncommon. Furthermore, even if some differences in treatment effects between men and women exist, perhaps typically they would not lead to different therapeutic decision-making. Our analysis of UpToDate information is consistent with this interpretation.

Our study has some limitations. First, we retrieved meta-analyses from only one database (PubMed). However, this set includes the large majority of medically relevant meta-analyses. We cannot generalize our findings to other fields like psychology or social sciences. Second, for some claims made in a potentially fallacious way, we could not exclude the possibility that a sex-treatment interaction might exist, because data were not provided to perform such an analysis. Nevertheless, the message that sex-treatment interactions are uncommon would not change. Third, we considerd p-values unadjusted for multiplicity, and often multiple subgroup analyses are performed. Thus even some sex-treatment interactions with statististical support may have been spurious. Fourth, statistically significant differences may sometimes reflect small differences in the absolute magnitude of treatment effects. This reinforces the message that clinically relevant sex-treatment interactions are scarce.

Subgroup analyses remain an area where much improvement is possible. When performing subgroups analysis, researchers should provide information to allow reproducing the results; and remain cautious in making strong claims [127], discussing the limitations and clinical implications (if any) of their findings. Given the current evidence, both statistically significant and clinically meaningful sex-treatment interactions seem rare.

## Data Availability

All data produced in the present study are available upon reasonable request to the authors

## AUTHORS CONTRIBUTIONS

**Lum Kastrati**-Conceptualized the study, screened the articles, evaluated claims, extracted the data, performed statistical analyses, checked UpToDate, wrote the manuscript and approved the final manuscript

**Sara Farina**-Screened the articles, evaluated claims, extracted the data, edited and approved the final manuscript

**Angelica-Vals Griz-** Evaluated claims, extracted data and checked for updated meta-analysis, edited and approved the final manuscript

**Hamidreza Raeisi-Dehkordi**, Evaluated claims, extracted data and checked for updated meta-analysis edited and approved the final manuscript

**Erand Llanaj** Evaluated claims, extracted data edited and approved the final manuscript

**Hugo G. Quezada-Pinedo** Evaluated claims, extracted data edited and approved the final manuscript

**Lia Bally** Conceptualized the study, edited and approved the final manuscript

**Taulant Muka** Conceptualized the study, edited and approved the final manuscript

**John P.A. Ioannidis** Conceptualized and supervised the study, performed statistical analyses, wrote the manuscript, edited and approved the final manuscript

---

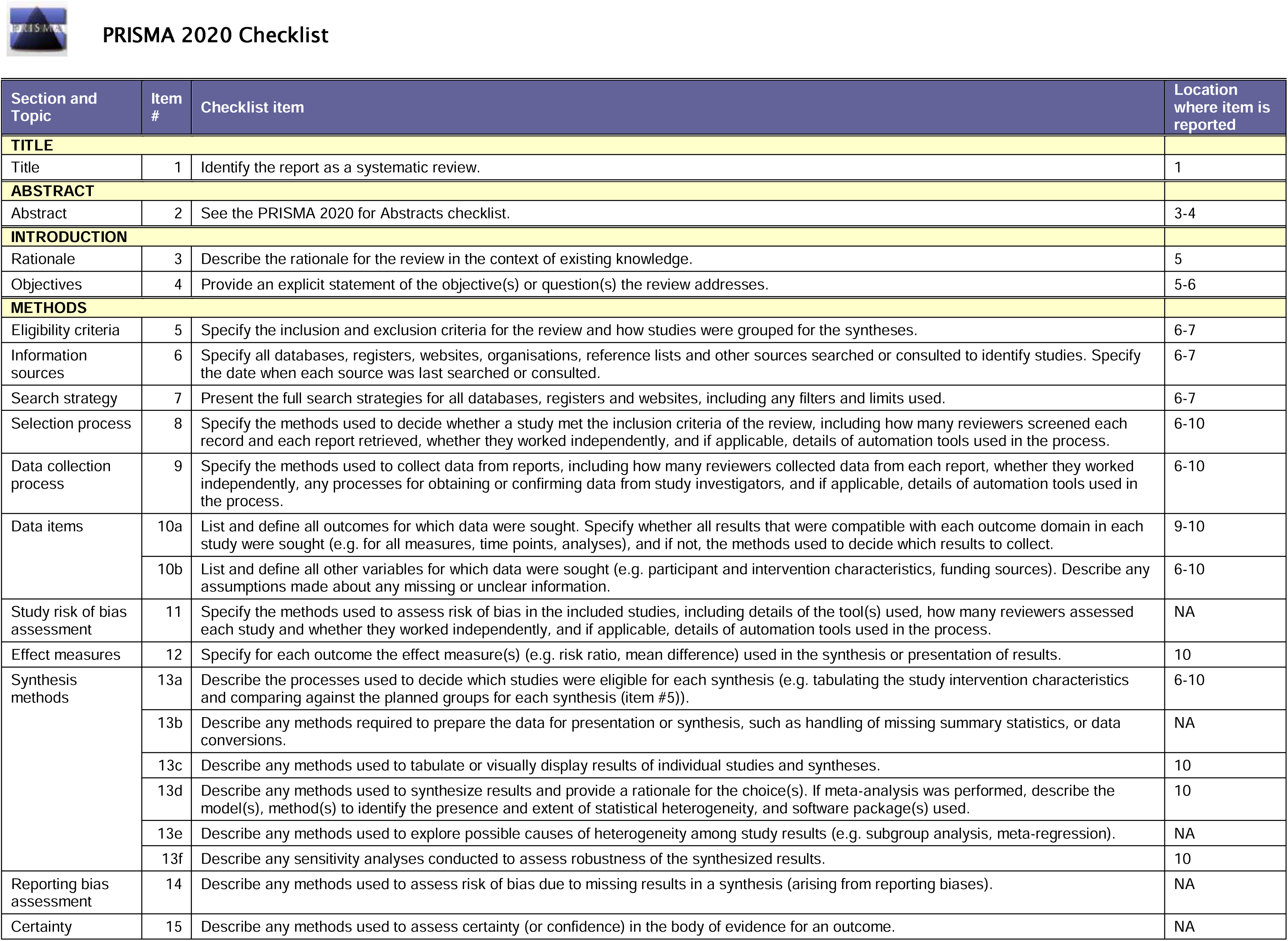
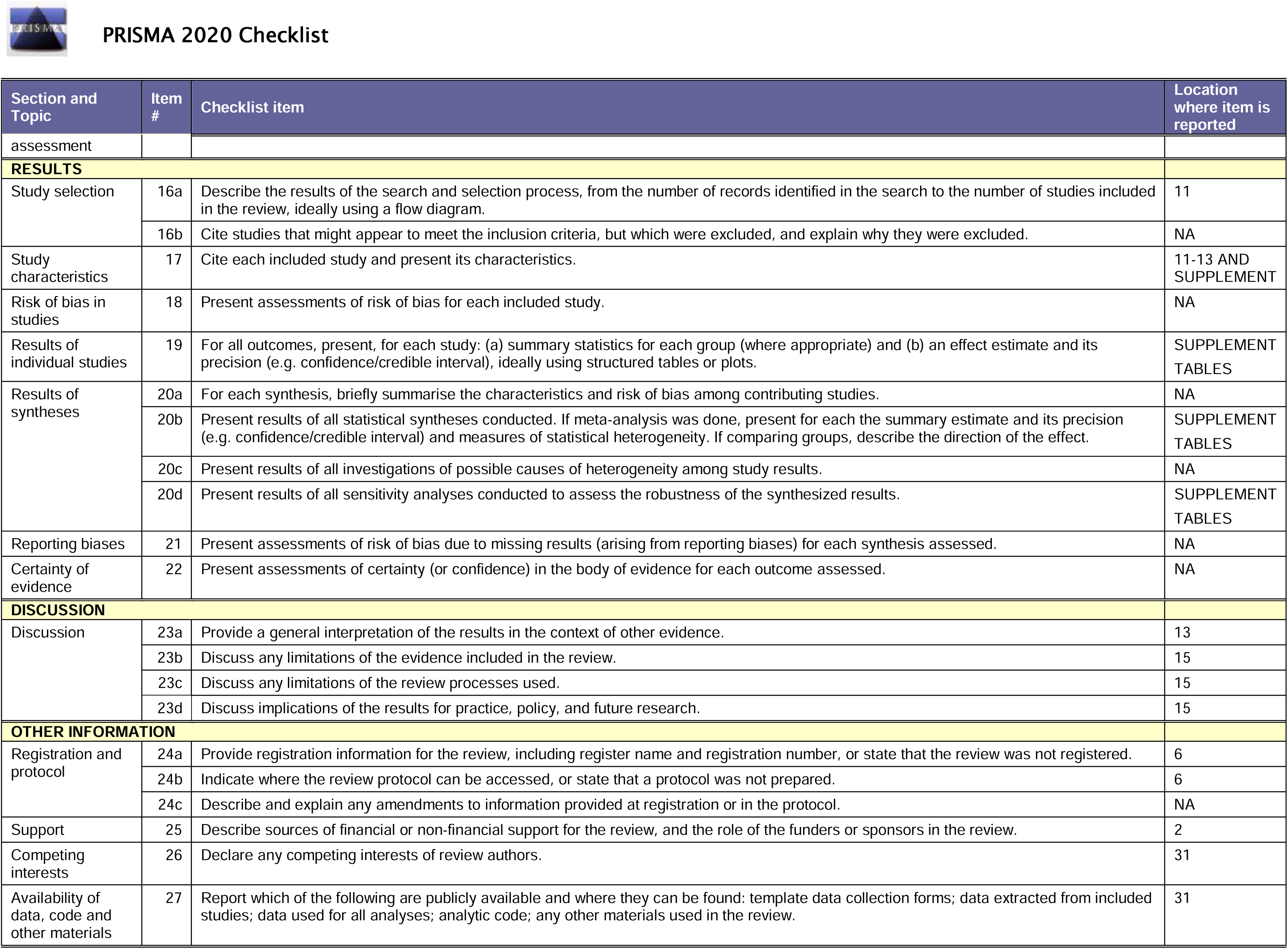
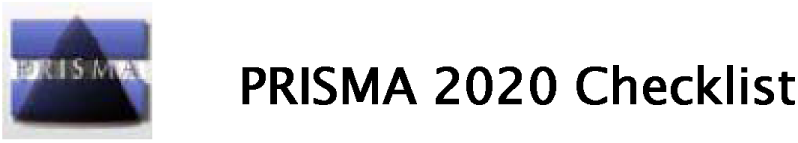

