## Supplementary Figures for "Evaluation of reported claims of sex-based differences in treatment effects across meta-analyses: A meta-research study"

**FIGURE S1.** Recalculation of subgroup analysis for X. Shu 2021 (random effects (D-L) top left, HKSJ top right, fixed effects bottom left) [1]

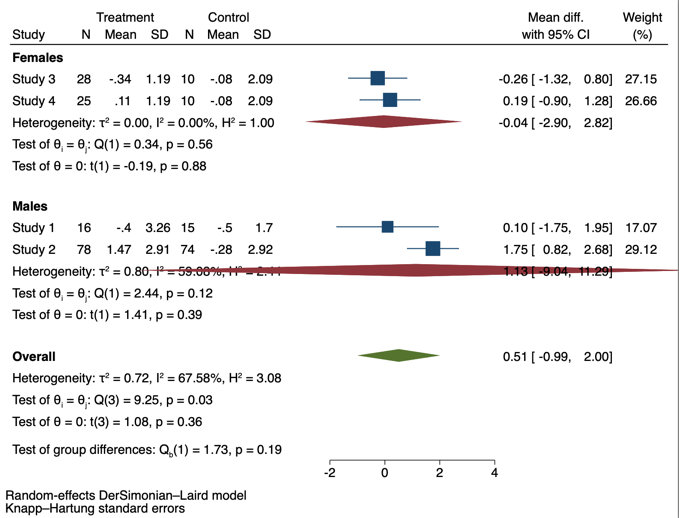

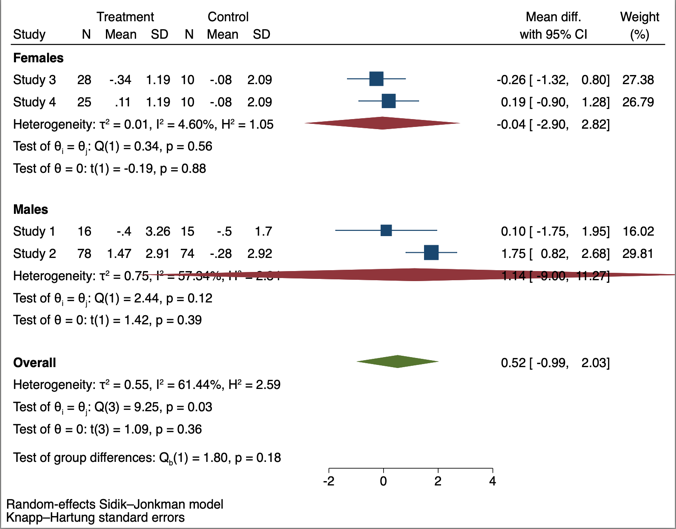

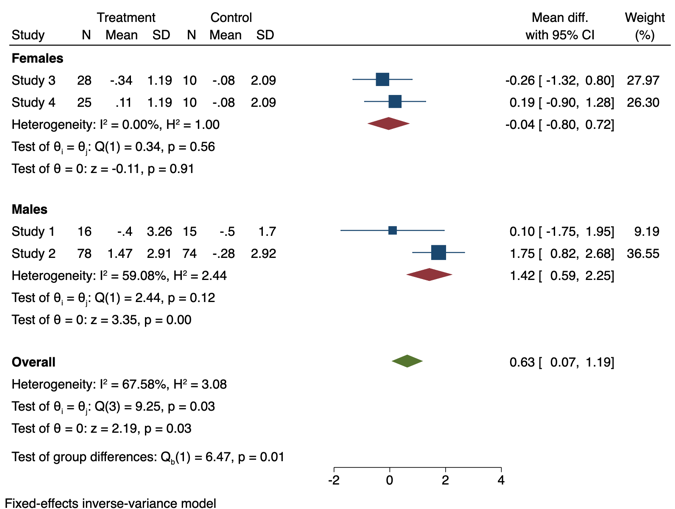

**FIGU1E S2a.** Recalculation of subgroup analysis for J. Zhao 2023 (random effects (D-L) top left, HKSJ top right, fixed effects bottom left) [2]

**Fasting plasma glucose**

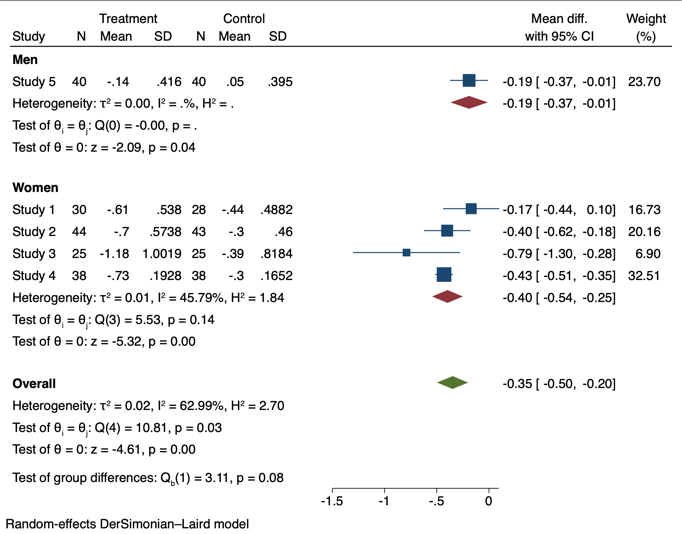

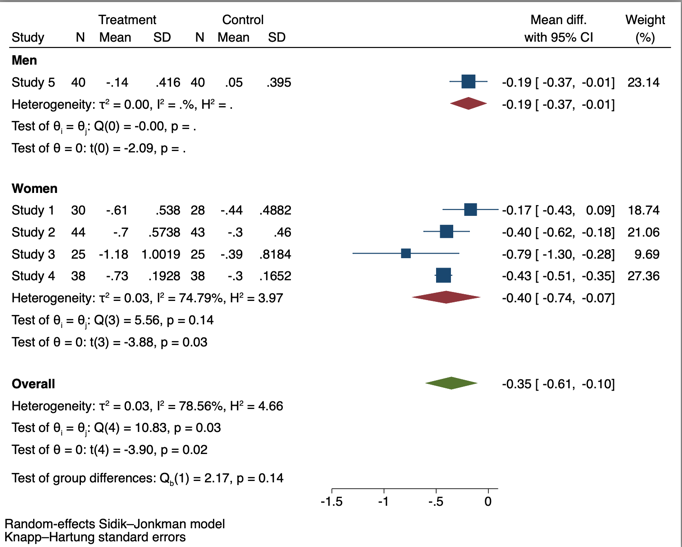

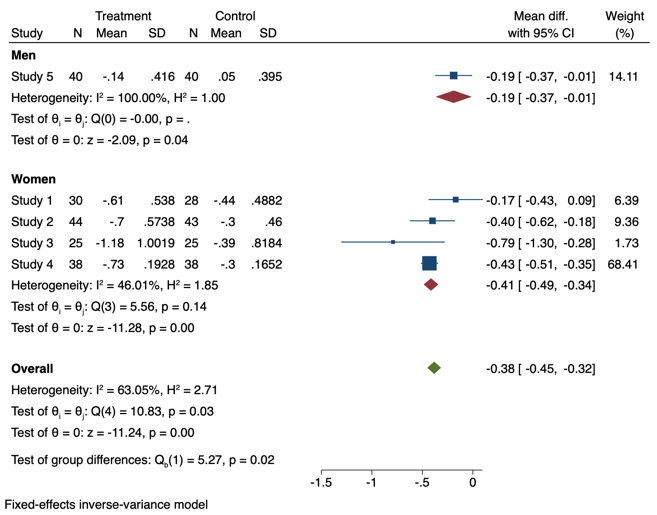

**FIGURE S2b.** Recalculation of subgroup analysis for J. Zhao 2023 (random effects (D-L) top left, HKSJ top right, fixed effects bottom left) [2]

**HOMA-IR( Insulin Resistance)**

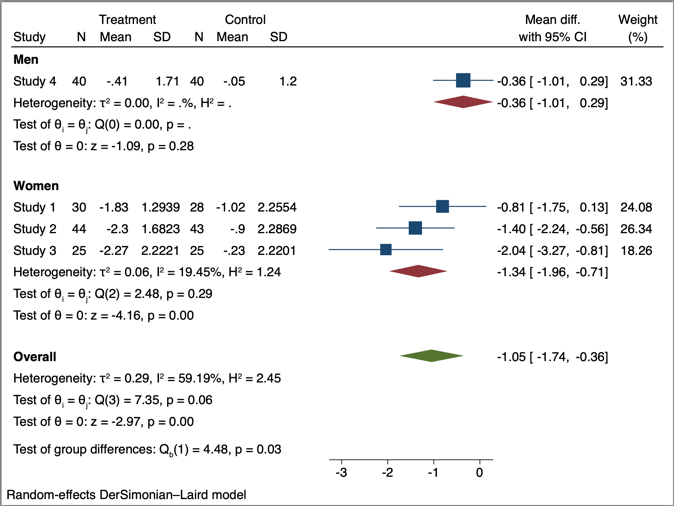

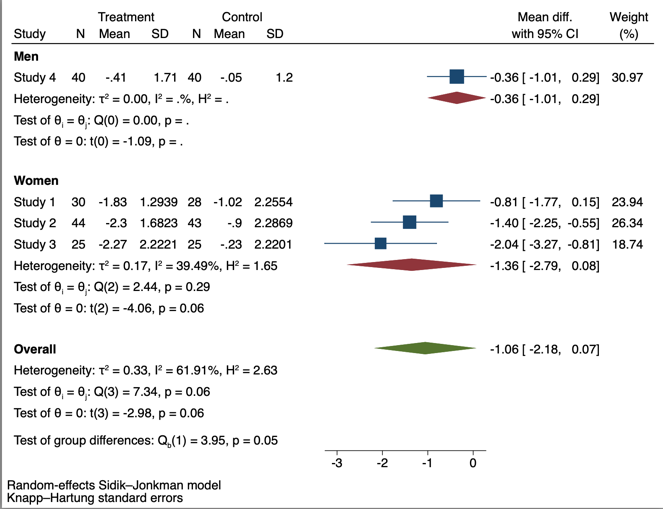

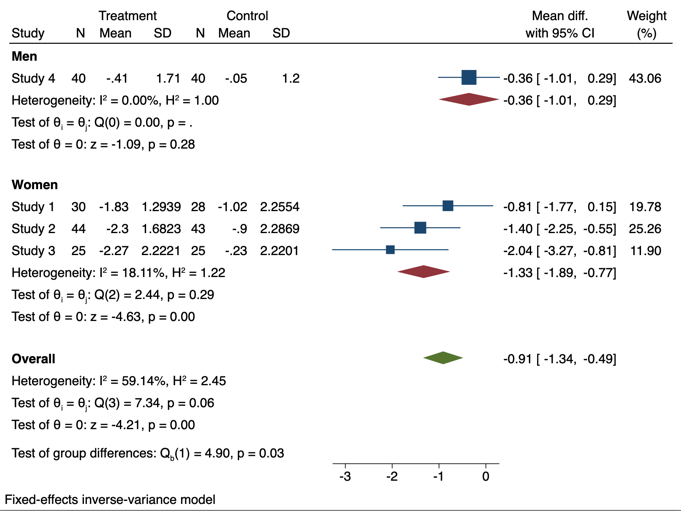

**FIGURE S3.** Recalculation of subgroup analysis for Chambers 2005 (random effects (D-L) top left, HKSJ top right, fixed effects bottom left) [3]

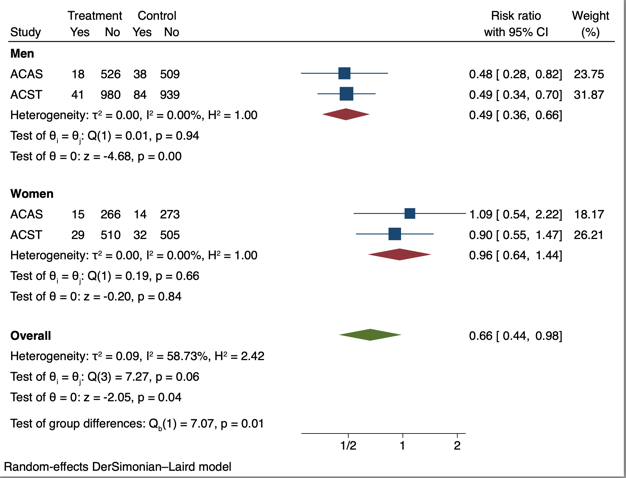

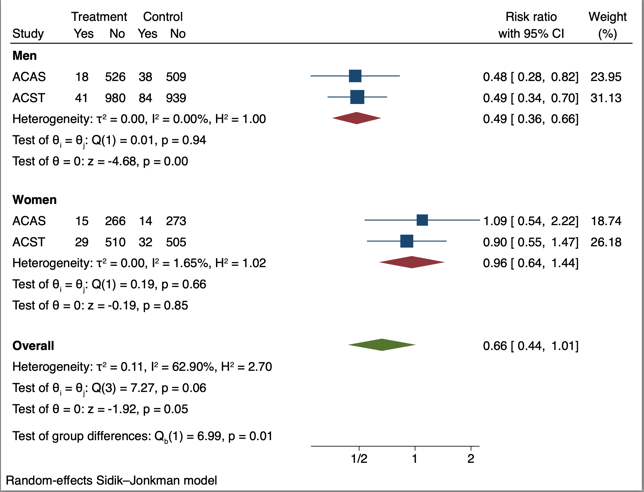

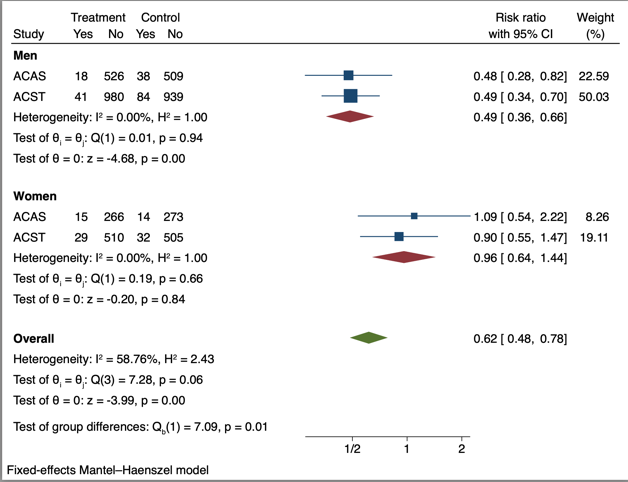

**FIGURE S4a.** Recalculation of subgroup analysis for M. Xie 2024 (random effects (D-L) top left, HKSJ top right, fixed effects bottom left) [4]

**Myocardial infarction**

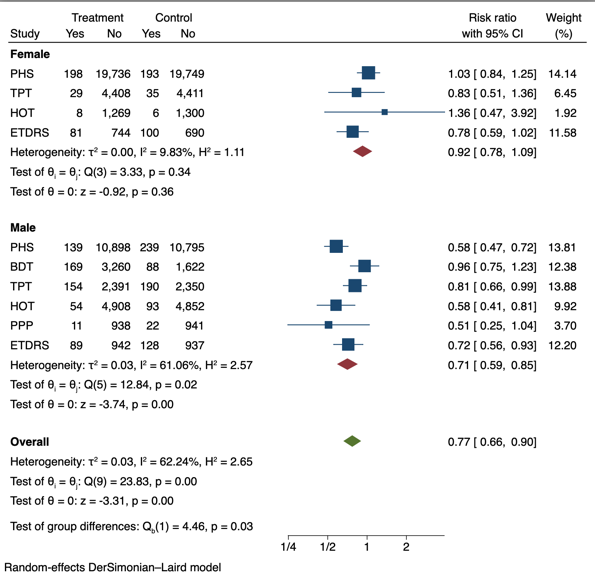

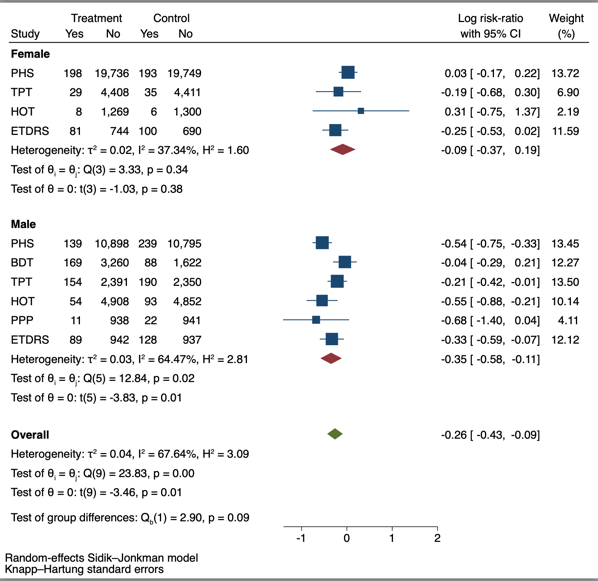

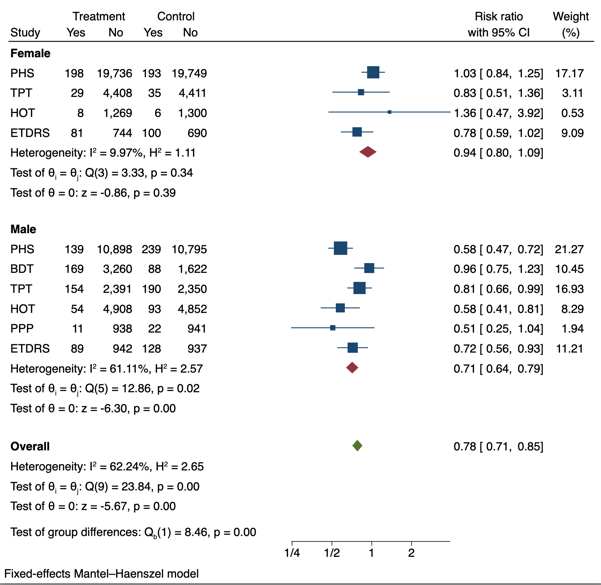

**FIGURE S4b.** Recalculation of subgroup analysis for M. Xie 2024 (random effects (D-L) top left, HKSJ top right, fixed effects bottom left) [4]

**Stroke**

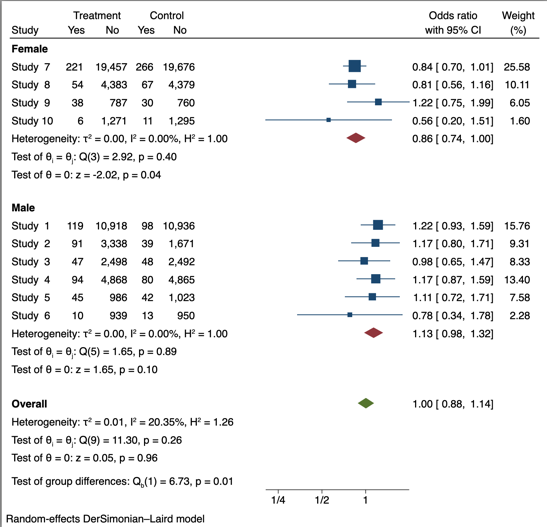

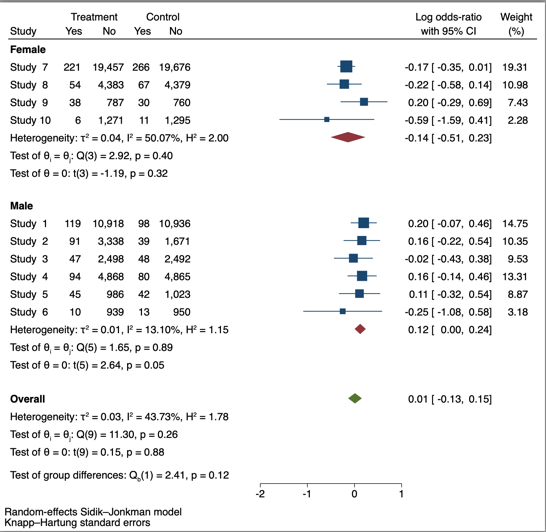

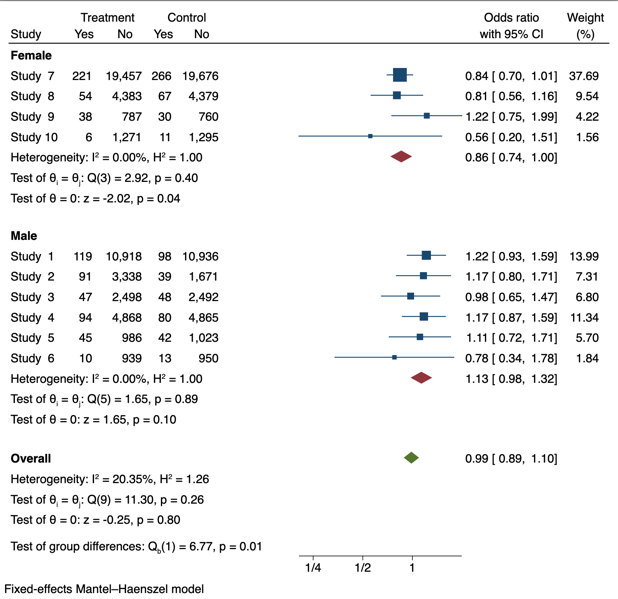

**FIGURE S5.** Recalculation of subgroup analysis for T. Rustagi 2015 (random effects left, fixed effects right)[5]

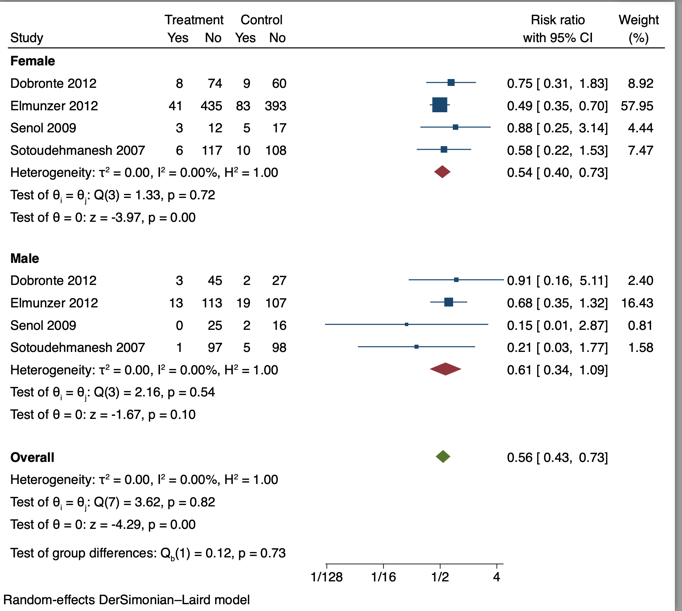

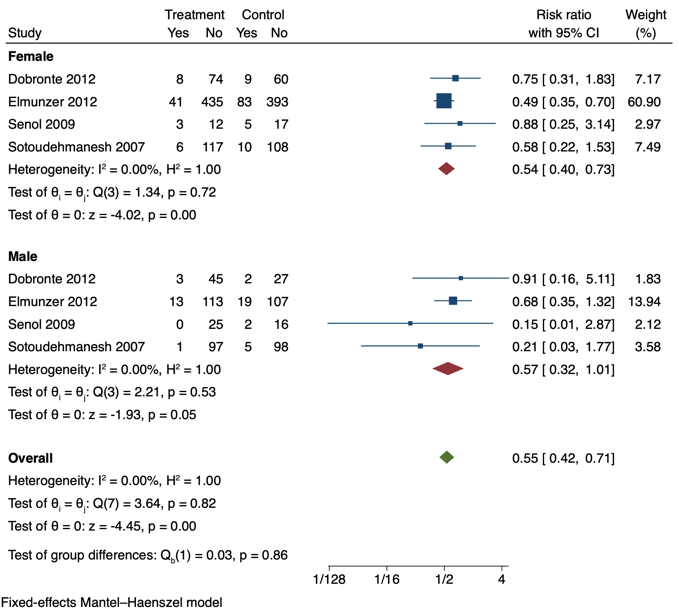

**FIGURE S6.** Recalculation of subgroup analysis for G. Gelbenegger 2019 (random effects left, fixed effects right)[6]

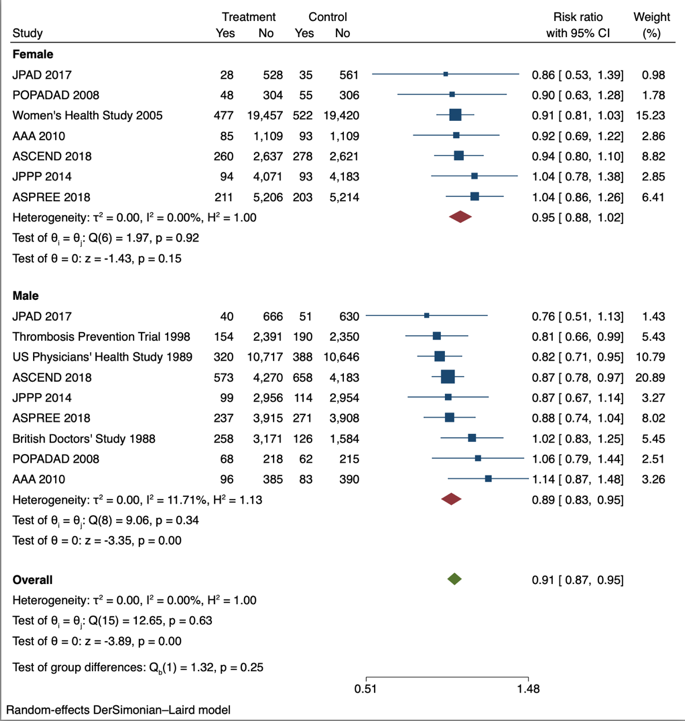

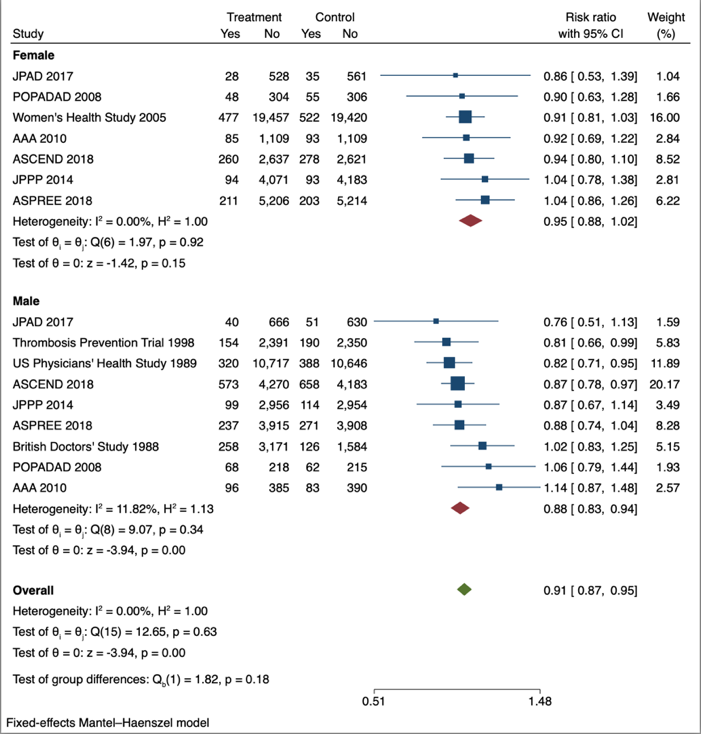

**FIGURE S7.** Recalculation of subgroup analysis for Z. Jiang 2022 (random effects left, fixed effects right)[7]

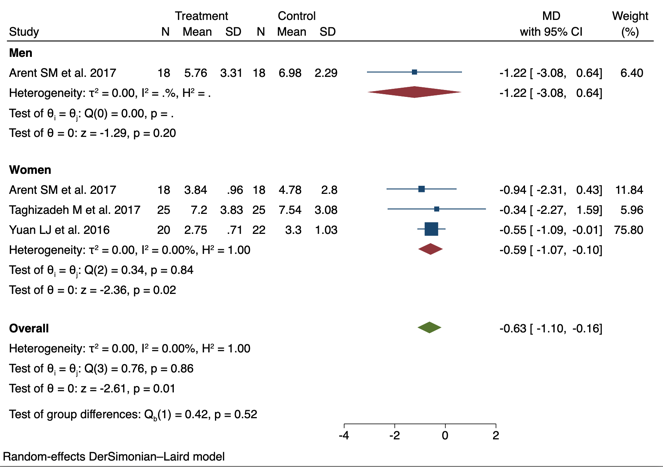

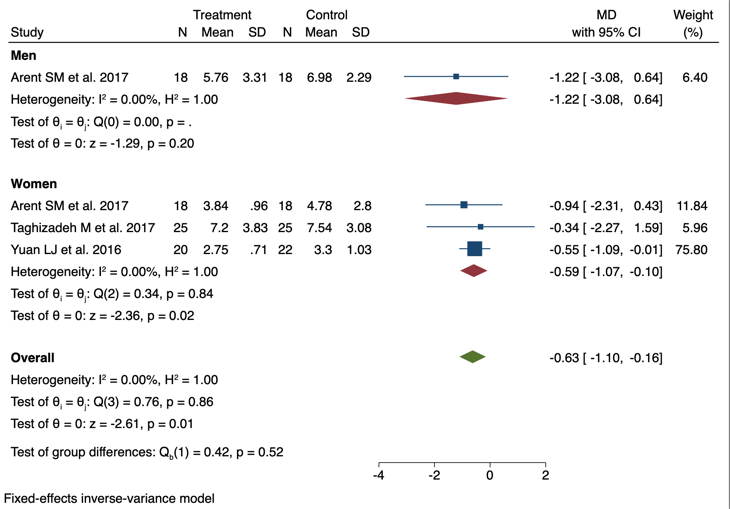

**FIGURE S8.** Recalculation of subgroup analysis for Hoffman 2022 (random effects left, fixed effects right)[8]

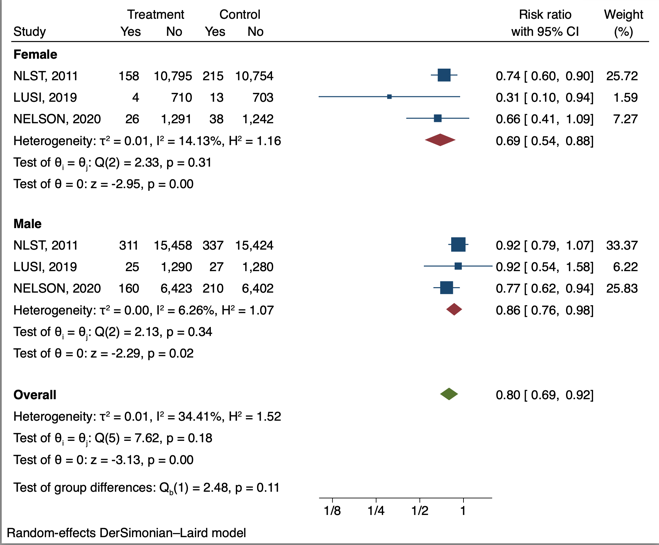

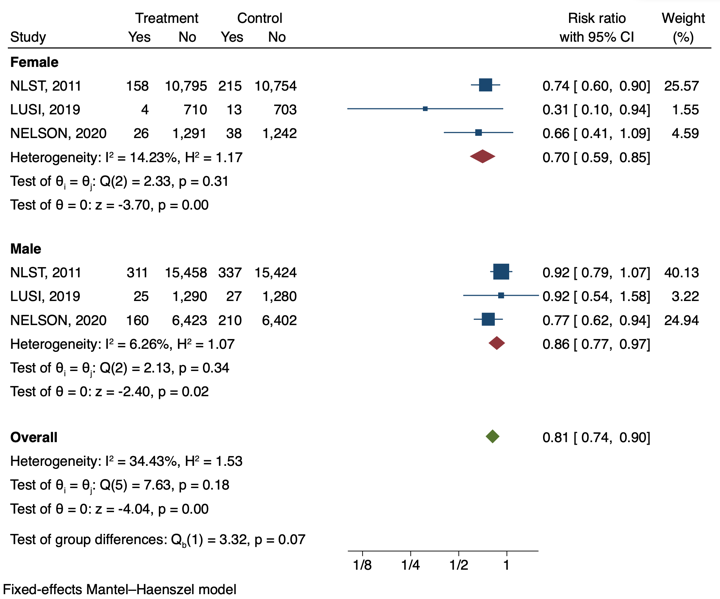

**FIGURE S9.** Recalculation of subgroup analysis for N. Amiri 2021 (random effects left, fixed effects right)[9]

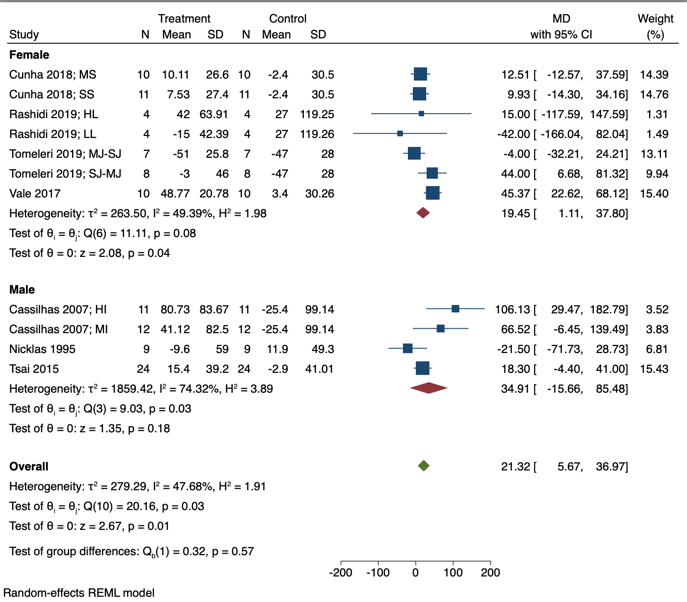

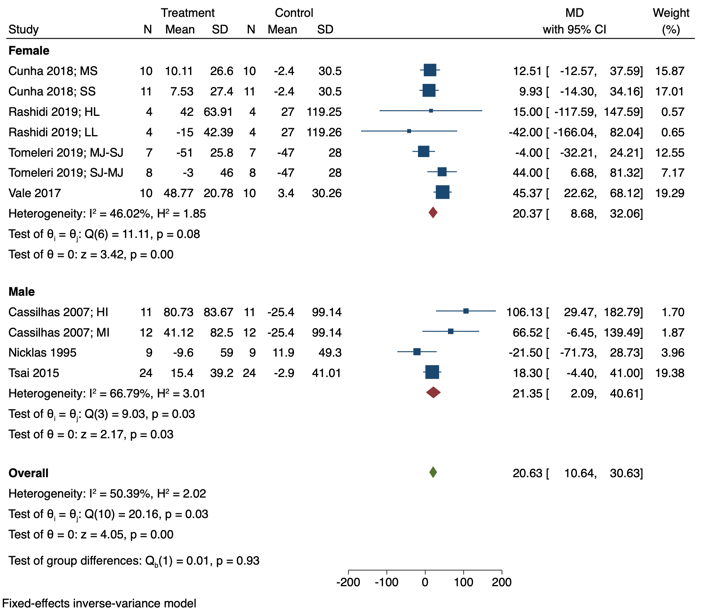
