## Supplementary Tables for "Evaluation of reported claims of sex-based differences in treatment effects across meta-analyses: A meta-research study"

**Table S1. Studies that mention in the abstract a plan to performe some sex-based subgroup analysis but reported no relevant claims.**

| Author, year | Title |
| --- | --- |
| M. Banerjee, 2023 | GLP-1 Receptor Agonists and Risk of Adverse Cerebrovascular Outcomes in Type 2 Diabetes: A Systematic Review and Meta-Analysis of Randomized Controlled Trials |
| K. Bourbeau, 2020 | The Combined Effect of Exercise and Behavioral Therapy for Depression and Anxiety: Systematic Review and Meta-Analysis |
| D. Breadner, 2020 | Efficacy and safety of ALK inhibitors in ALK-rearranged non-small cell lung cancer: A systematic review and meta-analysis |
| P. C. den Boer, 2005 | Paraprofessionals for anxiety and depressive disorders |
| D. Ghazvineh, 2022 | The Effect of Yoga on the Lipid Profile: A Systematic Review and Meta-Analysis of Randomized Clinical Trials |
| C. Heneghan, 2012 | Self-monitoring of oral anticoagulation: systematic review and meta-analysis of individual patient data |
| W. C. Jacobs, 2005 | Retention versus sacrifice of the posterior cruciate ligament in total knee replacement for treatment of osteoarthritis and rheumatoid arthritis |
| G. Jeet, 2007 | Community health workers for non-communicable diseases prevention and control in developing countries: Evidence and implications |
| D. A. Jolliffe, 2019 | Vitamin D to prevent exacerbations of COPD: systematic review and meta-analysis of individual participant data from randomised controlled trials |
| R. Menezes, 2017 | Impact of Flavonols on Cardiometabolic Biomarkers: A Meta-Analysis of Randomized Controlled Human Trials to Explore the Role of Inter-Individual Variability |
| J. Olin, 2001 | Hydergine for dementia |
| S. Shen, 2022 | Omega-3 Fatty Acid Supplementation and Coronary Heart Disease Risks: A Meta-Analysis of Randomized Controlled Clinical Trials |
| T. M. Winzenberg, 2010 | Vitamin D supplementation for improving bone mineral density in children |
| Y. Xie, 2021 | Effects of Exercise on Sleep Quality and Insomnia in Adults: A Systematic Review and Meta-Analysis of Randomized Controlled Trials |
| A. J. Clark, 2004 | Efficacy and safety of transdermal fentanyl and sustained-release oral morphine in patients with cancer and chronic non-cancer pain |
| D. A. Jolliffe, 2019 | Adjunctive vitamin D in tuberculosis treatment: meta-analysis of individual participant data |
| S. Leucht, 2022 | The response of subgroups of patients with schizophrenia to different antipsychotic drugs: a systematic review and meta-analysis |
| N. Schranz, 2013 | What is the effect of resistance training on the strength, body composition and psychosocial status of overweight and obese children and adolescents? A Systematic review and meta-analysis |
| N. G. Xunlin, 2020 | The effectiveness of mindfulness-based interventions among cancer patients and survivors: a systematic review and meta-analysis |
| Z. Ma, 2022 | Colchicine and coronary heart disease risks: A meta-analysis of randomized controlled clinical trials |

**Table S2. General information on the studies with negative claims**

| Author, year | Condition | Intervention | Control | Outcome | Claim |
| --- | --- | --- | --- | --- | --- |
| V. Arciero, 2023 | Cancer | Systemic oncology therapies (chemotherapy, targeted agents, immunotherapy) | Not specified | Overall survival (OS), Progression-free survival (PFS) | Female and male patients appear to derive comparable benefits from recently approved systemic oncology therapies." |
| O. Asbaghi, 2023 | Hypertension | Folic acid supplementation | Not specified | Blood pression reduction (Systolic blood pressure and diastolic blood pressure) | "Subgroup analysis showed that the results remained significant when baseline SBP was ≥120 mmHg, intervention duration was ≤6 weeks, intervention dose was ≥5 mg/d, in patients with CVD, males and females, and overweight participants, respectively. Furthermore, the changes observed in diastolic BP (DBP) (WMD: -0.24 mmHg; 95% CI: -0.37 to -0.10; *p* < 0.001) were also statistically significant. However, subgroup analysis showed that the results remained significant in subject with elevated DBP, long term duration of intervention (>6 weeks), low dose of folic acid (<5 mg/day), CVD patients, both sexes and male, and participants with normal BMI." |
| E. Ashford, 1998 | Migrain | Sumatriptan nasal spray (20 mg and 10 mg) | Placebo | Headache relief, adverse events | "No clinically significant differences in headache relief or adverse event rates were observed for any of the subgroups examined." |
| I. Bighelli, 2018 | Schizophrenia | Cognitive behavioural therapy (CBT) | Not specified | Overall symptoms reduction | "Blinding of outcome assessor, number of sessions, treatment duration, age and gender were not significant moderators of response." |
| H. A. Bischoff-Ferrari, 2004 | Falls in elderly individuals | Vitamin D supplementation | Calcium or placebo | Risk of falling | "Subgroup analyses suggested that the effect size was independent of calcium supplementation, type of vitamin D, duration of therapy, and sex, […]." |
| S. Burdett, 2015 | Early stage non small cell lung cancer (NSCLC) | Surgery plus adjuvant chemotherapy or surgery plus radiotherapy plus chemotherapy | Surgery alone or surgery plus radiotherapy | Overall survival (OS), Recurrence-free survival (RFS) | "There was little variation in effect according to the type of chemotherapy, other trial characteristics or patient subgroup, […]." |
| X. Cai, 2016 | Type 2 diabetes mellitus (T2DM) | Pedometer | No pedometer | Weight loss (BMI or weight) | "However, none of the following variables had a significant influence: step goal setting, baseline age, BMI, weight, sex distribution, […]." |
| Y. X. Cao, 2018 | Hypercholesterolemia | PCSK9 monoclonal antibodies (mAbs) | Placebo | Circulating high-sensitivity C reactive protein (hs-CRP) levels | "Meta-regression analyses suggested no significant linear correlation between baseline age (p=0.673), sex (p=0.645) and low-density lipoprotein cholesterol reduction (p=0.339)." |
| Y. X. Cao, 2018 | New-onset diabetes mellitus (NODM) | PCSK9 monoclonal antibodies (mAbs) | Different lipid-lowering therapy strategies or placebo | Risk of NODM | "Meta-regression analyses showed that risk of NODM was not associated with baseline age, baseline body mass index (BMI), proportion of men, treatment duration or percent LDL cholesterol reduction." |
| S. Chai, 2022 | Hyperglucagonemia in patients with type 2 diabetes mellitus (T2DM) | Dipeptidyl peptidase-4 (DPP4) inhibitors | Placebo or other oral antidiabetic drugs (OADs) | Reduction in circulating glucagon levels | "Analysis of subgroups revealed that study characteristics had no significant effect on results, such as study design (parallel group or crossover), number of patients, mean patient age, proportion of men, […] (all P for subgroup differences >0.05)" |
| F. T. Chen, 2020 | Executive function in older adults | Exercise training | No excercise training | Improvement in executive function | "No significant differences in effects were observed as a function of sex [Q(2) = 5.38, p > 0.05]." |
| L. Chen, 2023 | Type 2 diabetes mellitus (T2DM) | Sodium-glucose cotransporter 2 inhibitors (SGLT-2i) | Placebo or another antihyperglycemic as oral monotherapy | Urinary tract infections (UTI) incidence | "No significantly increased risk of UTI observed in subgroup analysis by gender. " |
| A. R. Coggan, 2021 | Muscle power | Dietary nitrate supplementation | Placebo | Changes in peak or maximal musle power | "Sub-group analyses revealed no significant differences due to subject age, sex, or test modality (i.e., small vs. large muscle mass exercise). " |
| M. Coylewright, 2014 | Chest pain, myocardial infarction, diabetes mellitus, osteoporosis | Decision aids (DAs) | Usual care | Knowledge transfer, decisional conflict, patient involvement in share decision making (SDM) | "These gains were largely consistent across sociodemographic patient groups, with DAs demonstrating similar efficacy when used with vulnerable patients such as the elderly and those with less income and less formal education. Differences in efficacy were found only in knowledge of risk in 1 subgroup, with greater efficacy among those with higher education (35% versus 18%; P=0.02)." |
| P. Cuijpers, 2018 | Depression | Psychotherapy | No psychotherapy | Effect on depression symptoms, using Beck Depression Inventory (BDI) or the Hamilton Rating Scale for Depression (HAM-D) | "A meta-regression analysis among low RoB studies showed that effect sizes found for studies among women, […], did not differ significantly from effect sizes from other studies. " |
| E. D'Andrea, 2020 | Major adverse cardiovascular events (MACE) | GLP-1 receptor agonists (GLP-1 RA) and SGLT-2 inhibitors (SGLT-2i) | Placebo or no active treatment | Risk of MACE in patients with type 2 diabetes | "Uncontrolled hypertension, obesity, gender, age and race did not appear to modify the effect of these drugs." |
| P. H. Decaup, 2023 | Mandibular cortical thickness | Environmental mechanical loading | Not Specified | Effect of environmental loading on mandibular cortical thickness | "Sex was found to be unrelated to cortical thickness pattern." |
| F. Dentali, 2015 | Stroke, systemic embolism and Venous Thromboembolism (VTE) | Direct Oral Anticoagulants (DOACs)/Non-vitamin K antagonist oral anticoagulants (NOACs) | Vitamin K antagonists or placebo | Efficacy: Prevention of stroke, systemic embolism, recurrent VTE, or VTE-related death; Safety: Occurrence of major and/or clinically relevant non-major bleeding | "No gender-related difference in the efficacy and safety of NOACs in patients with AF or acute VTE was found." |
| A. Drahman, 2023 | Post-Operative Urinary Retention (POUR) after Inguinal Hernia Repair (IHR) | Pre-operative alpha-blockers (e.g., Tamsulosin, Prazosin, Alfuzosin) | Placebo or no treatment | Risk of post-operative urinary retention (POUR) | "Gender did not affect the difference of incidence of POUR post-IHR despite pre-operative alpha-blockers (95% CI 0.62 (0.27, 1.44)), I2: 53%))." |
| N. El Hage Chehade, 2023 | Post-Endoscopic Retrograde Cholangiopancreatography Pancreatitis (PEP) | Combination of rectal nonsteroidal anti-inflammatory drugs (NSAID) and topical epinephrine (EI) | Rectal NSAID and normal saline (SI) | Risk of post-ERCP pancreatitis (PEP) | "The EI group did not demonstrate any significant benefit over SI group in preventing PEP, irrespective of gender or the epinephrine concentration used. The tests for subgroup differences were not statistically significant with P-values of 0.66 and 0.28, respectively." |
| I. Y. Elgendy, 2017 | Non-ST-elevation acute coronary syndrome (NSTE-ACS) | Routine invasive strategy | Selective invasive strategy | All-cause mortality risk | "There was no difference in treatment effect across various study-level covariates such as age, gender, diabetes, and positive troponin (all P for interaction >0.05)." |
| E. Esteve, 2015 | Groin injuries in sports | Specific groin-injury prevention programmes | Usual training/practice without specific prevention | Risk of groin injuries | "Subgroup analysis based on type of sports, gender and type of prevention programme showed similar non-significant estimates with RR ranging from 0.48 to 0.81." |
| A. J. Farmer, 2012 | Type 2 diabetes mellitus (T2DM) | Self monitoring of blood glucose levels | Clinical management without self monitoring | Reduction in HbA(1c) level | "The difference in HbA(1c) levels between groups was consistent across age, baseline HbA(1c) level, sex, […] were insufficient for interpretation." |
| O. Fasugba, 2017 | Catheter-associated urinary tract infections (CAUTIs) | Antiseptic cleaning before and during catheter use | No antiseptic cleaning | Risk of CAUTIs | "Subgroup analyses showed no difference in the incidence of CAUTIs in terms of country, setting, risk of bias, sex and frequency of administration." |
| J. Fulcher, 2015 | Major adverse cardiovascular events (MACE) | Intensive statin therapy | Less intensive statin therapy | Risk of major vascular events, major coronary events, stroke, coronary revascularisation, mortality, adverse events | "In men and women at an equivalent risk of cardiovascular disease, statin therapy is of similar effectiveness for the prevention of major vascular events." |
| G. M. Gager, 2021 | Heart failure (HF) | SGLT2 inhibitors | Placebo or glimepiride | Hospitalization for HF, cardiovascular mortality, all-cause mortality | "The effect of SGLT2 inhibitors on the primary endpoint was independent of underlying diabetes mellitus, age, sex, BMI, renal function, and HF type." |
| L. Gijsbers, 2016 | Heart rate | Potassium supplementation | Placebo | Effect on HR | "Stratified analyses yielded no significant effects of potassium intake on HR in subgroups" |
| M. Goyal, 2016 | Acute ischemic stroke | Endovascular thrombectomy | Standard medical care | Disability on modified Rankin Scale (mRS) at 90 days | "Subgroup analysis of the primary endpoint showed no heterogeneity of treatment effect across prespecified subgroups for reduced disability (pinteraction=0.43)." |
| J. P. Greving, 2019 | Noncardioembolic stroke or transient ischemic attack | Antiplatelet agents (Aspirin/dipyridamole combination, Clopidogrel, Aspirin/clopidogrel combination, aspirin) | Antiplatelet agents (Aspirin/dipyridamole combination, Clopidogrel, Aspirin/clopidogrel combination, aspirin) | Serious vascular events (nonfatal stroke, nonfatal myocardial infarction, or vascular death), Major bleeding, Net clinical benefit (serious vascular event or major bleeding) | "Subgroup analyses [age, sex, ...] showed no heterogeneity of treatment effectiveness across prespecified subgroups." |
| M. Guo, 2018 | Stroke | SGLT2 inhibitors | Placebo or standard care | Stroke risk in patients with type 2 diabetes | " Subgroup analyses showed that RR values were not affected by gender, age, diabetes duration, BMI or HbA1C levels" |
| T. Guo, 2021 | Anxiety disorders in children and adolescents | Individual cognitive behavior therapy (I-CBT) | Group cognitive behavior therapy (G-CBT) | Efficacy (mean change in anxiety symptom scores) at post-treatment | "However, the findings were not materially different from those of the efficacy subgroup analysis of number of treatment sessions, parental involvement, male/female sex, and number of participants." |
| F. Haghighatdoost, 2017 | Serum adiponectin concentration | Green tea | Placebo | Change in serum adiponectin levels | "Subgroup analyses based on sex, type of intervention, continent, and body mass index (BMI) could not explain the sources of heterogeneity." |
| A. H. Hai, 2021 | Substance use and related consequences in adults of color | Culturally adapted interventions (CAIs) | No CAIs | Substance use reduction | "Moderator analysis showed that CAIs’ effects might not vary significantly by treatment model, dose, country, follow-up assessment timing, participant age, or gender/sex." |
| N. N. Hariharan, 2022 | Embolic stroke of undetermined source (ESUS) | Direct oral anticoagulation (DOAC) (dabigatran or rivaroxaban) | Antiplatelet therapy (aspirin) | Recurrent stroke, major bleeding, clinically relevant non-major bleeding (CRNB) | "Prespecified subgroup analysis demonstrated consistent results according to age and sex." |
| A. C. Hogwood, 2023 | Excercise performance | Nitrate (NO3-) supplementation (e.g., beetroot juice) + exercise training | Placebo + exercise training | Longer-term exercise training responses (VO2peak, Time to Exhaustion (TTE)) | "No significant moderators were revealed on either outcome." |
| H. Hosseini, 2020 | Type 2 diabetes mellitus (T2DM) | Resveratrol supplementation | Placebo | C-reactive protein (CRP) level | "Random-effects meta-regression did not indicate any significant association of CRP level with potential confounders including resveratrol dose, duration of treatment, age and gender of type 2 diabetic patients." |
| Z. Huang, 2020 | Solid malignant tumors | SNHG12 expression | Not available | Overall survival (OS) | "[…], while there was no significant correlation between SNHG12 expression and gender." |
| K. C. Hung, 2021 | Quality of recovery | Intravenous lidocaine during the perioperative period | Placebo | Postoperative quality of recovery (QoR-40), intraoperative opioid consumption, risk of chronic postsurgical pain (CPSP) | "Subgroup analysis demonstrated no significant impact of the type of surgery, age, gender, […]." |
| K. C. Hung, 2022 | Hypoxemia during gastrointestinal endoscopic procedures (GEPs) under sedation | High Flow Nasal Oxygenation (HFNO) | Standard care without HFNO | Risk of hypoxemia | "Subgroup analysis focusing on risk of hypoxemia showed no significant subgroup effects, indicating consistent benefits of HFNO in different clinical settings." |
| N. J. Ives, 2017 | High-risk malignant melanoma | Interferon-α (IFN-α) adjuvant therapy | No IFN-α adjuvant therapy | Event-free survival (EFS), Overall survival (OS) | "There was no evidence that the benefit of IFN-α differed depending on dose or duration of treatment, or by age, gender, […]. " |
| S. Jamshidi, 2022 | Inflammatory markers (TNF-α, IL-6) status | Whey protein (WP) supplementation | Carbohydrate and other types of proteins | TNF-α and IL-6 status | "Results from subgroup analysis based on health status, study duration, WP dosage and sex, expressed no favorable effect of WP on TNF-α and IL-6 levels." |
| B. C. Johnston, 2018 | Clostridium difficile infection (CDI) | Probiotic prophylaxis | Placebo or no treatment (standard care) | Risk of CDI, adverse events | "Age, sex, hospitalization status, and high-risk antibiotic exposure did not [increase the odds of CDI]." |
| D. Kabbani, 2023 | COVID-19 infection requiring hospitalization | Immune-based therapy | No immune-based therapy | Risk of secondary infections | "A meta-regression revealed no impact of age, sex, or mechanical ventilation on the effect of immune-based therapies on risk of infection." |
| K. Khatri, 2023 | Osteoporosis fractures | Calcium and/or vitamin D supplementation | No treatment/placebo | Risk of fractures (hip, vertebral, any other fracture) | "Further subgroup analysis of the results did not vary with the dosage of calcium or vitamin D, dietary calcium intake, sex, or serum 25-hydroxyvitamin D levels." |
| S. Kuznia, 2023 | Cancer | Daily or bolus vitamin D3 supplementation | Placebo | Cancer mortality, all-cause survival, cancer-specific survival | "The IPD were used to test effect modification by age, sex, […] but no statistically significant findings were obtained in meta-analyses of all trials." |
| C. K. Lee, 2018 | Advanced non-small cell lung cancer (NSCLC) | Checkpoint inhibitors (nivolumab, pembrolizumab, or atezolizumab) | Docetaxel | Overall survival (OS) | "The relative treatment benefits were similar according to […] sex (male [HR, 0.69] vs female [HR, 0.70]; interaction, P = .82)." |
| L. L. Lee, 2021 | Hypertension | Walking as a physical activity intervention | No intervention | Effects on blood pressure (SBP and DBP) and heart rate (HR) | "Moderate- and low-certainty evidence suggests walking may reduce DBP and heart rate for all ages and both sexes." |
| C. X. Li, 2023 | Type 2 diabetes mellitus (T2DM) | SGLT-2 inhibitors | Placebo | Risk of reproductive tract infections (RTIs) | "Compared to placebo, a significantly higher risk of RTIs was observed with canagliflozin, ertugliflozin, empagliflozin, remogliflozin, dapagliflozin, and sotagliflozin, but not with luseogliflozin and ipragliflozin, regardless of gender." |
| J. Li, 2023 | Cancer | Sintilimab combinations (with chemotherapy or targeted therapy) | Single treatment (chemotherapy or targeted therapy) | Progression-free survival (PFS) | "Subgroup analyses suggested that the sintilimab-chemotherapy group exhibited a superior PFS benefit than the chemotherapy alone group regardless of age, gender, […]." |
| L. F. Li, 2021 | STEMI myocardial infarction with multivessel disease | Complete revascularization | Culprit-only revascularization | Major adverse cardiac events (MACE) | "All of the subgroup effects based on the 5 factors of interest (i.e., sex, age, […]) were not statistically significant (Psubgroup ranged from 0.198 to 0.556)." |
| L. Lin, 2022 | Neonatal growth | Early macronutrient supplements | No supplementation | Cognitive impairment and metabolic risk | "In subgroup analysis, supplementation increased the height z-score in male toddlers (aMD 0.20[0.02, 0.37], *p* = 0.03; 10 trials, *n* = 595) but not in females, and no significant sex interaction was observed (*p* = 0.21)." |
| K. H. Liu, 2019 | Adult androgenic alopecia (AGA) | Low-level laser therapy (LLLT) | Sham devices | Increase in hair density | "LLLT represents a potentially effective treatment for AGA in both male and female." |
| X. Liu, 2019 | Atherosclerosis | Atorvastatin | Placebo or blank treatment | Effects on circulating adiponectin | "Results of univariate meta-regression analyses showed that study characteristics including number of patients, mean age, proportion of male patients, […] did not significantly affect the outcome (p all > 0.05)." |
| Z. Liu, 2021 | Acute kidney injury (AKI) after cardiac surgery | remote ischemic preconditioning (RIPC) | No RIPC | Risk of AKI | "Meta-regression and subgroup analyses indicated that study characteristics, including [...] gender, […], did not significantly affect the outcome of AKI." |
| K. M. Livingstone, 2016 | Overweight/obesity | Dietary, physical activity, or drug-based interventions | No intervention | Effect of the FTO genotype on weight loss after interventions | "Differential changes in body mass index, body weight, and waist circumference by FTO genotype did not differ by intervention type, intervention length, ethnicity, sample size, sex, and baseline body mass index and age category." |
| S. Lueangarun, 2021 | Pattern Hair Loss (PHL) | FDA-approved home-use LLLT devices | Sham devices | Increase in hair density | "The subgroup analysis demonstrated the increased hair growth in male and female subjects with both comb- and helmet-type devices." |
| F. Lv, 2023 | Fractures | PCSK9 inhibitors (alirocumab, evolocumab, bococizumab, inclisiran) | Placebo | Risk of major osteoporotic fracture, hip fracture, osteoporotic non-vertebral fracture, and total fracture | "No significant associations were detected in any of the sensitivity analyses and subgroup analyses stratified by the type of PCSK9i, follow-up duration, age, sex, sample size, and patient profile." |
| A. Mahmoudi, 2022 | Depression, Quality of Life, Muscle Strength | Aerobic training, resistance training, combined training | No training | Effect on depression symptoms, quality of life, and muscle strength in healthy people aged 60 or more. | "Subgroup analysis revealed that depression decreased significantly when aerobic training (p = .000) and resistance training (p = .003) were applied, and for studies including both genders (p = .000) or men subjects (p = .002)." |
| B. Man, 2021 | Arterial stiffness | Soy isoflavone supplementation | Placebo | Arterial stiffness reduction | "Subgroup analyses showed no difference between treatment effects for intervention duration (< 6 weeks vs. ≥ 6 weeks) or gender (women only vs. men only vs. combined)." |
| E. Mannucci, 2020 | Major adverse cardiovascular events (MACE) | GLP-1 receptor agonists | Placebo | Risk of MACE | "The meta-analyses of patient subgroups showed a significant reduction in MACE with GLP-1RAs, irrespective of gender, advanced age and obesity. " |
| J. W. Martha, 2022 | Contrast-induced nephropathy (CIN) | Pentoxifylline | Placebo or no intervention | Mortality | "Meta-regression analysis showed that the association between pentoxifylline and mortality was not affected by age (p=0.994), gender (reference: male, p=0.562), […]." |
| A. Mousa, 2018 | Type 2 diabetes mellitus (T2DM) | Vitamin D supplementation | Placebo or usual care | Reduction of chronic low-grade inflammation (CRP, TNF-α, ESR, leptin, adiponectin, IL-6, and E-selectin) in patients with type 2 diabetes | "In meta-regression and subgroup analyses, age, sex, […] did not alter the results." |
| S. M. Mousavi, 2018 | Obesity | Nigella sativa supplementation | Placebo | Weight loss (BMI or weight) | "Subgroup analysis by the intervention type (I(2)=0.0%), participants' gender (I(2)=0.0%),[…]. Subgroup analysis based on study duration (I(2)=0.0%), participants' gender (females: I(2)=0.0% & both genders: I(2)=20.9%), an age (I(2)=35.9%) disappeared the heterogeneity (BMI)." |
| S. Nath, 2018 | Lumbar puncture complications | Atraumatic needles | Conventional needles | Postdural-puncture headache incidence | "Prespecified subgroup analyses of postdural-puncture headache revealed no interactions between needle type and patient age, sex, […]." |
| J. Park, 2021 | Obesity in children | Child-centered information and communication technology (ICT) interventions | Traditional obesity interventions | BMI, body weight, BMI z-score, waist circumference, and percentage body fat | "Subgroup analyses revealed that the effect of the intervention was statistically significant for the following: [...], intervention involving both boys and girls (WMD=-1.30, 95% CI -2.14 to -0.46), […]" |
| F. Passiglia, 2021 | Lung cancer | Computed tomography lung screening (CTLS) | No screening (NS) or Chest x-ray (CXR) | Lung cancer mortality | "The analysis of lung cancer-related mortality by sex revealed nonsignificant differences between men and women (P = .21; I-squared = 33.6%)." |
| K. Prokopidis, 2023 | Memory | Creatine supplementation | Placebo | Memory performance | "Creatine dose (≈ 2.2-20 g/d), duration of intervention (5 days to 24 weeks), sex, or geographical origin did not influence the findings." |
| T. Qin, 2015 | Colorectal cancer | Folic acid supplementation | Placebo or no intervention | Colorectal cancer risk | "Moreover, no statistical effect was identified in further subgroup analyses stratified by ethnicity, gender, body mass index (BMI) and potential confounding factors." |
| M. Qiu, 2021 | Atrial fibrillation following percutaneous coronary intervention (PCI) | Double antithrombotic therapy (DAT) | Triple antithrombotic therapy (TAT) | Clinically significant bleeding, major adverse cardiac events (MACE) | "This effect [on cinically significant bleeding] of DAT was observed in most subgroups of interest (HR ranged from 0.54 to 0.69), and was consistent across various subgroups defined by each of the 5 factors of interest (Psubgroup ranged from 0.290 to 0.794). [...] This effect [on MACE] of DAT was observed in all subgroups of interest (all 95% CIs of HRs were across 1.0), and was consistent across various subgroups defined by each of the 5 factors of interest (Psubgroup ranged from 0.308 to 0.828)." |
| N. Radkhah, 2021 | Cardiovascular disease (CVD) | Vitamin D supplementation | Placebo | Apolipoproteins A1 and B100 levels | "No significant effects of vitamin D on Apo A1 and Apo B100 levels after subgroup analysis by mean age, gender, study population, dosage and duration of study." |
| H. Reinink, 2021 | Space-occupying hemispheric infarction | Surgical decompression | Conservative treatment | Mortality, favorable outcomes (modified Rankin Scale [mRS] score ≤3) | "Surgical decompression was associated with a decreased chance of death (adjusted odds ratio, 0.16; 95% CI, 0.10-0.24) and increased chance of a favorable outcome (adjusted odds ratio, 2.95; 95% CI, 1.55-5.60), without evidence of heterogeneity of treatment effect across any of the prespecified subgroups [age, sex, ...]." |
| Y. M. Roman, 2019 | Overweight/obesity | Intermittent dieting (regular or intensified) | Continuous dieting | Weight, body fat, lean mass, waist circumference, hip circumference, energy expenditure | "Subgroup effects by time to follow-up, gender, per-protocol versus intention-to-treat, enforced exercise, and diabetes were similar to main analyses." |
| E. Rosenfeld, 2009 | Growth of term and preterm children | Formula milk with LC-PUFAs (DHA and AA) | Formula milk without LC-PUFAs | Weight for age, length for age, head circumference for age, BMI for age at 18 months | "Multivariate regression analyses including the possible confounders, sex, […], as well as interaction terms showed no significant effects of LC-PUFA supplementation on any z-score." |
| J. M. Ross, 2021 | HIV and Tuberculosis | Isoniazid preventive therapy + antiretroviral therapy (ART) | ART without isoniazid | Risk of incident tuberculosis and all-cause mortality | "[…], but there was no significant difference in the benefit of isoniazid preventive therapy with ART by sex, baseline CD4 count, or results of tuberculin skin test or IGRAs." |
| J. Runhaar, 2017 | Hip or knee osteoarthritis | Oral glucosamine | Placebo | Effect on pain and function | "Glucosamine was also no better than placebo among the predefined subgroups [pain, age, sex and BMI]." |
| A. W. Scholtz, 2022 | Central and/or peripheral vestibular vertigo | Cinnarizine 20 mg and dimenhydrinate 40 mg | Placebo or cinnarizine, dimenhydrinate, betahistine dimesylate, betahistine dihydrochloride. | Reduction of mean vertigo score (MVS) | "Subgroup analyses with respect to age groups (< 65 years/≥ 65 years) and sex showed no significant differences in efficacy within any of the treatment groups." |
| A. Schweizer, 2010 | Cardiovascular and cerebrovascular (CCV) safety of vildagliptin | Vildagliptin (50 mg qd or 50 mg bid) | Placebo or other treatments | Risk of CCV events (acute coronary syndrome, transient ischaemic attack, stroke, CCV death) | "The results were consistent across subgroups defined by age, gender and CV risk status […]" |
| P. Shekarchizadeh-Esfahani, 2021 | Effect of cinnamon supplementation on liver enzymes | Cinnamon supplementation | Placebo | Liver enzymes (AST, ALT, ALP) levels | "Subgroup analyses showed that the effect of cinnamon supplementation on ALT was significant at the dosages of <1500 mg/day (Hedges’s: -0.61; 95 % CI: -1.11, -0.10; P = 0.002), [...] and in trials conducted of both gender (Hedges’s: -0.72; 95 % CI: -1.45, -0.01; P = 0.04)." |
| Y. Shi, 2021 | Non-squamous non-small cell lung cancer (NSCLC) | Immune checkpoint inhibitors (ICIs) (e.g., pembrolizumab, atezolizumab) + platinum-doublet chemotherapy | Platinum-doublet chemotherapy | Progression-free survival (PFS) | "Subgroups including PD-1 status [high (>50%), intermediate (1-49%), and negative (<1%) expression], sex (male and female), […] were all associated with better PFS." |
| F. Shiraseb, 2022 | Hypertension | Oral l-arginine supplementation | Placebo | Systolic blood pressure (SBP) and diastolic blood pressure (DBP) | "Subgroup analysis showed significant reductions in SBP and DBP regardless of baseline BP category (normotensive, hypertensive), study duration (≤24 d, >24 d), sex (female, male), […]" |
| D. B. Shrestha, 2021 | Heart failure | SGLT-2 inhibitors | Placebo | Mortality, cardiovascular mortality, hospitalization for heart failure (HHF) | "In addition, a benefit was seen for composite outcome HHF or mortality and considering subgrouping based on diabetes status, gender, and age groups." |
| M. G. Sim, 2023 | Cardiovascular disease | Calcium supplementation | Not specified | Myocardial infarction, total stroke, heart failure admission, cardiovascular mortality, all-cause mortality | "Subgroup analysis focusing on calcium monotherapy/calcium co-therapy with vitamin D, female sex, follow-up duration, and geographical region did not affect the findings." |
| A. F. Smelt, 2018 | Haematological parameters in elderly | Vitamin B12 or folic acid supplementation | Placebo | Hematological parameters changes | "The effects did not differ by sex or by age group." |
| M. Sohn, 2023 | Type 2 diabetes mellitus (T2DM) | SGLT-2 inhibitors (SGLT-2i) or GLP-1 receptor agonists (GLP-1RA) | Not specified | Incidence of 3-point major adverse cardiovascular events (3P-MACE) | "Other factors such as age, sex, BMI, HbA1c, and preexisting CVD or HF did not affect the efficacy of either treatment on the ARR or RRR of 3P-MACE." |
| A. G. Solimando, 2022 | Hepatocellular carcinoma (HCC) | Second-line treatments (Regorafenib, Cabozantinib, Ramucirumab, Brivanib, S-1, Axitinib, Pembrolizumab) | Placebo | Overall survival (OS), progression-free survival (PFS), and drug withdrawal due to adverse events | "None of the compared drugs deemed undoubtedly superior after having performed a patients' stratification [Subgroup stratification considered gender, …]." |
| R. S. Taylor, 2019 | Heart failure (HF) | Exercise-based cardiac rehabilitation (ExCR) | No exercise | Mortality (all-cause and HF-specific), hospitalisation (all-cause and HF-specific), exercise capacity, health-related quality of life (HRQoL) | "No strong evidence for differential intervention effects across patient characteristics was found for any outcomes. [age, sex, ...]" |
| R. A. Tejada, 2017 | HPV-related lesions | HPV quadrivalent vaccine | Not specified | Risk of anogenital warts (AGW) | "The quadrivalent vaccine reduced the risk of AGW by 62% (RR: 0.38, 95%CI: 0.32-0.45, I2: 0%) in ITT analysis and by 95% (RR: 0.05, 95%CI: 0.01-0.25, I2: 66%) in PP analysis. Subgroup analyses of studies in women or with low risk of bias provided similar results." |
| C. Wagner, 2022 | COVID-19 | Systemic corticosteroids | Standard care and placebo | All-cause mortality, clinical improvement, clinical worsening, serious adverse events, adverse events, infections | "We conducted the following subgroup analyses to explore equity-related factors: sex, age [...], ethnicity [...]. Except for age and ethnicity, no evidence for differences could be identified." |
| D. Wang, 2015 | Cardiovascular disease (CVD) | Fibrates | Placebo or no treatment | Composite outcome of non-fatal stroke, non-fatal myocardial infarction (MI), vascular death | "Subgroup analyses showed the benefit of fibrates on the primary composite outcome to be consistent irrespective of age, gender, and diabetes mellitus." |
| D. Wang, 2019 | Exercise-induced Fatigue | Laser therapy | Placebo | Lactate levels, repetitions | "The results of subgroup analyses indicated no significant differences between the laser therapy and placebo for lactate and repetitions when stratified by study design, mean age, gender, and study quality." |
| Y. Y. Wang, 2018 | Major Depressive Disorder (MDD) | Mindfulness-based interventions (MBIs) | Treatment as usual (TAU) | Reduction in depressive symptoms | "Subgroup analyses revealed that positive benefits of MBIs was associated with studies that had treatment as usual (TAU) control group, Chinese participants, open label design, no gender predominance […]." |
| M. A. Wewege, 2022 | Body Composition | Full-body resistance training | No exercise | Body fat percentage and body fat mass | "Measurement type was a significant moderator in body fat percentage and body fat mass, but sex was not." |
| Y. Xu, 2021 | Non-small cell lung cancer (NSCLC) | Immune checkpoint inhibitors (ICIs) | Non-ICI therapy | Overall survival (OS) | "While sex, histological type, PS 0 or 1, KRAS mutation status and region were not associated with the efficacy of ICIs. " |
| F. Yang, 2020 | Advanced cancer | Immune checkpoint inhibitors (ICIs) | Non-ICI therapy | Overall survival (OS) | "No significant difference of relative benefit from immunotherapy over control therapy was found in patients of different sex (P = .25, I2 = 19.02%), […]." |
| D. Yin, 2021 | Heart failure (HF) | Gliflozins | Placebo | Composite outcome of cardiovascular death, or hospitalization for HF | "The benefit of gliflozins in HF patients was not affected by the other 10 factors (Psubgroup≥ .123) [status of type 2 diabetes, sex, ...]." |
| J. Zhang, 2019 | Coronary stenting | Dual therapy (DT) | Triple therapy | Safety (relative bleeding events), and efficacy (major adverse cardiovascular events) | "In subgroup analyses, results were similar by sex, bleeding risk, and stent type" |
| J. G. Zhao, 2017 | Osteoporosis-related fractures | Calcium, vitamin D, or combined calcium and vitamin D supplements | Placebo or no treatment | Risk of hip fracture, nonvertebral fracture, vertebral fracture, and total fracture. | "Subgroup analyses showed that these results were generally consistent regardless of the calcium or vitamin D dose, sex, fracture history, dietary calcium intake, and baseline serum 25-hydroxyvitamin D concentration." |
| J. Mytton, 2006 | Aggressive behavior in children | School-based violence prevention programs for aggressive or at-risk children | No intervention | Aggressive behaviour reduction, school and agency responses to acts of aggression, or risk of violent injuries | "Benefits can be achieved in both primary and secondary school age groups and in both mixed sex groups and boys-only groups." |
| J. D. Chalmers, 2019 | Bronchiectasis | Long-term macrolide treatment | Placebo | Frequency of exacerbations requiring treatment with antibiotics | "Effect estimates in prespecified subgroup analyses revealed a reduced frequency of exacerbations in all prespecified subgroups [age, sex, ...]" |

**Table S3**. General information of analyses with sex-based differences claims that have statistical support at p<0.05

| **Author** | **Condition** | **Intervention** | **Control** | **Outcome** | **Abstract Claim** | **Genuine Claim Outcome** | **P-Value** | **Protocol availability** | **Prespecified subgroup analysis** |
| --- | --- | --- | --- | --- | --- | --- | --- | --- | --- |
| **Z. Q. Li, 2020** | Non-small cell lung carcinoma | PD-1/PD-L1 | Platinum-based chemotherapy | Progression free survival and overall survival | In patients subjected to the combined treatment regimen, we observed significant differences in Progression free Survival among groups stratified by PD-L1 expression (p < 0.001), immune drug type (p = 0.029), gender (p = 0.014) and liver metastasis (p = 0.035) and Overall survival among groups stratified by immune drug type (p < 0.001), gender (P = 0.001) and smoking status (P = 0.041). | Progression free survival | 0.014 | No | / |
|  |  |  |  |  |  | Overall survival | 0.001 |  |  |
| **A. Kazemi, 2018** | HIV | Probiotics | Placebo | CD4 cell count | Subanalysis showed that CD4 counts increased significantly after supplementation in studies with female participants (6.015; 95% CI [2.576, 9.454]; p = .001) while it decreased in studies concerning male participants (−43.69; 95% CI [−57.867, −29.522]; p < .001), | CD4 count | <0.001 | No | / |
| **H. C. Jodal, 2019** | Colorectal cancer | Faecal blood testing | Sigmoidoscopy/colonoscopy | Cancer incidence and mortality | Sigmoidoscopy had a greater effect in men, for both colorectal cancer incidence (women: RR 0.86; 95% CI 0.81 to 0.92, men: RR 0.75, 95% CI 0.71 to 0.79), and mortality (women: RR 0.85; 95% CI 0.71 to 0.96, men: RR 0.67; 95% CI 0.61 to 0.75) (moderate certainty). | Colorectal cancer incidence Mortality | 0.001  0.015 | Yes | Yes |
| **L. Gu, 2022** | Gastro-esophageal cancer | PD-1/PD-L1 | Placebo/other therapy | Overall survival | Overall survival of all subgroups containing male patients was significantly improved compared with chemotherapy, unlike that of female patients. | Overall survival | 0.001 | Yes | Yes |
| **B. R. Chambers, 2005** | Asymptomatic carotid stenosis | Carotid endarterectomy | Medication | Perioperative stroke or death or subsequent carotid stroke | CEA appeared more beneficial in men than in women and more beneficial in younger patients than in older patients although the data for age effect were inconclusive. | Perioperative stroke or death or subsequent carotid stroke | 0.008 | Yes | Yes |
| **M. Boulanger, 2015** | Myocardial infarction | Carotid angioplasty and stenting | Carotid Endarterectomy | Myocardial infarction | Only the effect of sex differed between CAS and CEA with men being at lower risk of MI than women after CAS, whereas there was no difference between after CEA (Pint=0.01). | Myocardial Infarction | 0.01 | No | // |
| **J. Yu, 2023** | Cardiovascular disease | Any breakfast before 10:00, | Water | Cardiovascular outcomes | Subgroup analysis also revealed potential factors that may affect the outcomes, for example, the physiological condition of participants, duration, gender, and type of breakfast. | Fasting blood glucose | <0.001 | No | / |
| **Zhao et al., 2023** | Healthy subjects | Berberine | Placebo | FPG & HOMA-IR | Effects on fasting glucose (FPG) and insulin resistance (HOMA-IR) showed potential differences by sex, with larger reductions in women than in men. | FPG | 0.049 | Yes | Yes |
|  |  |  |  |  |  | HOMA-IR | 0.03 |  |  |
| **X. Shu, 2019** | Healthy subjects | Pravastatin | Placebo | Changes of circulating adiponectin | Subgroup analyses confirmed that pravastatin significantly increased adiponectin in studies of males (WMD =1.41 µg/mL; P=0.008), but not in those of females (WMD =-0.04 µg/mL; P=0.94). | Adiponectin | 0.03 | No | / |
| **G. G. Soon, 2012** | HIV | Antiretroviral therapy | Different antiretroviral therapy | Not specified | However, the corresponding subgroup analyses appear to show several statistically significant gender differences favoring males | Response rate | 0.001 | No | / |
| **L. Wang, 2019** | Diabetes | Inulin-type fructans | Different types of sugars | FBG, HbA1c, FINS, and HOMA-IR | Moreover, subgroup analyses suggested that the effects of glycemic control were significantly influenced by the sex of the subjects. | Fasting insulin | <0.001 | Yes | No |
|  |  |  |  |  |  | Insulin resistance | 0.003 |  |  |
| **D. S. Inauen, 2023** | Dentistry | Orthodontic treatment with fixed appliances on one or both jaws | Any comparoson | Patient reported pain at different timepoints | Non-consistent effects were seen for patient age, sex, irregularity, or analgesic use. | Pain at day 5 | 0.03 | No | / |
| **N. M. A. d'Unienville, 2021** | Healthy subjects | Nitrates or polyphenol | No treatment | Exercise performance | No effect of nitrate or polyphenol consumption was found in females. | Endurance | 0.03 | Yes | No |
| **S. Rastgoo, 2023** | Healthy or unhealthy adults | Conjugated linoleic acid | Placebo/control diet | Adiponectin and leptin | A subgroup analysis found that CLA supplementation reduced adiponectin and leptin in women. | Leptin | 0.0048 | Yes | No |
| **A. Hadi, 2019** | Adult population | Purslane | Placebo/nutrition | Blood lipids and glucose | Categorization based on gender showed that purslane was more effective in improving FBG, TC and LDL-C in females compared with males. | Fasting blood glucose | 0.008 | Yes | Yes |
|  |  |  |  |  |  | Total cholesterol | 0.02 |  |  |
| **T Jiang, 2018** | Non-small cell lung cancer | PD-1/PD-L1-inhibitor | Chemotherapy or placebo | Overall survival or Progression free survival | Subgroup analysis showed that anti-PD-1/PD-L1 monotherapy could markedly improve OS in elderly patients (HR, 0.69; P < .001), female patients (HR, 0.70; P < .001), never-smoking patients (HR, 0.73; P = .001), and patients with a histology of squamous cell carcinoma (HR, 0.67; P < .001), but not PFS in the elderly and female patient groups. | Progression Free Survival | 0.03 | No | / |
| **Y. H. Wang, 2022** | Healthy subjects | Physical activity interventions | Different levels of activity | Circulating brain-derived neurotrophic factor | Via subgroup analysis, studies of long-term PE with larger sample sizes, female participants, participants older than 60 years, and aerobic exercise contributed to a more pronounced improvement on BDNF levels than that found when all studies were combined. | Brain derived neutrophic factor | 0.038 | No | / |
| **Zhou et al., 2017** | Diffuse large B-cell lymphoma | Rituximab maintenance therapy | Observation | Event-free survival (EFS) | A subgroup analysis suggested that male patients may benefit from rituximab maintenance therapy with a better EFS (HR = 0.53, 95% CI: 0.34-0.82-), while this advantage was not observed in female patients (HR = 0.99, 95% CI: 0.64-1.52). | Event- free survival | 0.048 | No | / |
| **Loffredo, 2016** | Venous thromboembolism | New oral anticoagulants | Standard of care | Not specified | The subgroup analysis showed a significant gender difference in incidence of major bleedings and clinically relevant minor bleedings only for Edoxaban (RR: 0.52; 95% CI: 0.42–0.64; p < 0.001). | Major bleedings and clinically relevant minor bleedings only | <0.001 | No | / |
| **Shiraseb, 2022** | Blood pressure | L-Arginine | Placebo | Blood pressure | l-Arginine supplementation also appears to decrease DBP more effectively in females than in male. | Dyastolic Blood Pressure | 0.001 | Yes | No |
| **Xie, 2014** | Cardiovascular Disease | Aspirin | Placebo | Cardiovascular events | The effects of aspirin therapy varied by sex and diabetes status. | Myocardial Infarction | 0.02 | Yes | Yes |
|  |  |  |  |  |  | Stroke | 0.01 |  |  |

# ‘

### Table S4. General information of analyses with fallacious sex-based differences claims

| **Author, year** | **Condition** | **Intervention** | **Control** | **Outcome of interest** | **Fallacious claims** | **Abstract Claim** | **Fallacy category** | **P-value** |
| --- | --- | --- | --- | --- | --- | --- | --- | --- |
| **T. Rustagi, 2015** | Post–Endoscopic Retrograde Cholangiopancreatography Pancreatitis | Nonsteroidal anti-inflammatory drugs (NSAIDs) | Placebo | Post–Endoscopic Retrograde Cholangiopancreatography Pancreatitis | primary | Nonsteroidal anti-inflammatory drugs were effective in both young and old females (RR, NSAIDs did not significantly reduce the risk of PEP in men (RR, 0.61; 95% CI, 0.34–1.09) | 2.0 | 0.73 |
| **Z. U. A. Asad, 2918** | Atrial Fibrilation | Catheter ablation | Medical therapy | All-cause mortality | Composite outcome of death, stroke, major bleeding, cardiac hospitalization, and cardiac arrest with catheter ablation vs medical therapy | Interestingly, men (HR, 0.60; CI, 0.46–0.78; P=0.0002) but not women (HR, 0.97; CI, 0.62–1.53; P=0.91) derived a significant benefit from CA compared with MT (Figure 8). | 2.0 | 0.687 |
| **I. B. Riaz, 2013** | Cryptogenic stroke | Transcatheter patent foramen ovale closure | medical therapy | death, stroke and transient-ischemic attack (TIA). | primary | Significant benefit for male for the primary outcome | 2.0 | 0.146 |
| **A. H. Malik, 2020** | Percutaneous Coronary Intervention | Dual antiplatelet therapy | Monotherapy | Major bleeding, ischaemic events and all-cause and cardiovascular mortality | Major bleeding | For the outcome of major bleeding the results favor the use of P2Y12 monotherapy in the subgroups of age, sex, diabetes, chronic kidney disease, acute coronary syndrome, and multivessel disease. | 2.0 | 0.143 |
| **W. Liu, 2023** | Non-small cell lung cancer | Pembrolizumab | Chemotherapy | Overall survival | Overall survival | Pembrolizumab significantly enhanced OS for the first line treatment in male (HR 0.74; CI 95%, 0.65–0.83; p < 0.00001), but not in female individuals (HR 0.57; CI 95%, 0.31–1.06; p = 0.08) compared with chemotherapy (Fig. S1B | 2.0 | 0.415 |
| **W. Liu, 2022** | Non-small cell lung cancer | Atezolizumab | Chemotherapy | Overall survival | Overall survival | Subgroup analysis revealed that male individuals, those with non-squamous NSCLC, those with PS 1, active or previous smokers, and those with wild-type EGFR, TC3 or IC3, and TC1/2/3 or IC1/2/3 achieved OS benefit from atezolizumab treatment not related to the treatment line and treatment regimen. | 2.0 | 0.862 |
| **Kunutsor, 2017** | Cardiovascular disease | Aspirin | Control | MACE | MACE | stratified analysis by sex showed that aspirin significantly reduced the risk of MACE in men 0.79 (95% CI 0.64 to 0.98; P = 0.033) but not in women 0.95 (95% CI 0.77 to 1.16; P = 0.591; P value for meta-regression = 0.437). | 2.0 | / |
| **A. Hadi, 2019** | Adult population | Purslane | Placebo/nutrition | Blood lipids and glucose | TG | Categorization based on gender showed that purs-lane was more effective in improving FBG, TC and LDL‐C in females compared withmales. | 2.0 | TG= 0.29 |
| **A. Grassadonia, 2018** | Advanced Cancer | Imune checkpoints inhibitorsq | Treatment/placebo | Not specified | Overall survival or progression free survival | 1. the anti-CTLA-4 treatment was effective in men (HR 0.77, 95% CI 0.63–0.94, p = 0.012) (Figure 4A), but did not reach significance in women (HR 0.89, 95% CI 0.76–1.05, p = 0.162)  2. Meta-analysis using the random-effects model revealed a significant improvement in PFS in men (HR 0.67, 95% CI 0.55–0.80, p < 0.001, I2 73%) (Figure 5A), but not in women (HR 0.77, 95% CI 0.57–1.05, p = 0.100, I2 63%) | 2.0 | 1=0.27  2=0.45 |
| **G. Gelbenegger, 2019** | Cardiovascular disease | Aspirin | Control | All-cause mortality | MACE | 1. Aspirin did not reduce the RR of IS in men (RR 1.02; 95% CI, 0.72–1.44; p = 0.93; I2 = 55%). In women, however, aspirin reduced IS by 23% (RR 0.77; 95% CI, 0.63–0.94; p = 0.010) compared to control as reported in one study.  2. Aspirin in men was associated with a RRR of MACE of 11% (RR 0.89; 95% CI, 0.83–0.95; p = 0.0008; I2 = 12%; Fig. 5) compared with controls. In women, aspirin did not significantly reduce the risk of MACE (RR 0.95; 95% CI, 0.88–1.02; p = 0.16; I2 = 0%; Fig. 5) compared with controls. | 2.0 | 0.25 |
| **J. Gao and L. Yu, 2023** | - | Strength–endurance sequence | Endurance-strength sequence | VO2max and lower limb strength | Lower limb strength | the S-E sequence of females was more conducive to the growth of lower limb strength than of males (p = 0.017). | 2.0 | 0.2 |
| **V. Formica, 2021** | Gastric carcinoma | PD-1/PD-L1 inhibitors | Other control | Not specified | Survival | Interestingly our meta-analysis identified a possible interaction between ICI and gender (Fig. 5). Such a differential efficacy of immunotherapy in favour of the male gender has been observed in other disease settings (e.g. lung cancer) (Conforti et al., 2018). | 2.0 | 0.15 |
| **T Jiang, 2018** | Non-small cell lung cancer | PD-1/PD-L1-inhibitor | Chemotherapy or placebo | Overall survival or Progressiong Free Survival | Overall survival | Subgroup analysis showed that anti-PD-1/PD-L1 monotherapy could markedly improve OS in elderly patients (HR, 0.69; P < .001), female patients (HR, 0.70; P < .001), never-smoking patients (HR, 0.73; P = .001), and patients with a histology of squamous cell carcinoma (HR, 0.67; P < .001), but not PFS in the elderly and female patient groups. Notably, PD-1/PD-L1 inhibitors cannot prolong both OS (HR, 0.76; P = .390) and PFS (HR, 0.74; P = .210) in patients with central nervous system (CNS) metastasis, whereas patients without CNS metastasis could benefit from anti-PD-1/PD-L1 monotherapy on OS (HR, 0.71; P < .001). | 2.0 | 0.88 |
| **Z. Jiang, 2022** | Metabolic syndrome | Capsaicin | Placebo | Lipid profile | Triglyceride Levels | The results revealed that CAP decreased TG levels in women (WMD = −0.59, 95% CI: −1.07 to −0.10) | 2.0 | 0.52 |
| **Xie et al., 2020** | Healthy population | DHEA | Placebo | Serum IGF-1 levels | Serum IGF-1 levels | DHEA supplementation increases IGF-1 levels, especially in women and older subjects | 2.0 | 0.053 |
| **J. Sheng, 2015** | Non-small cell lung cancer | Antiangiogenic agents + taxanes-containing chemotherapy | Taxanes-containing chemotherapy | Overall survival | Overall survival | Other clinical factors directing significant OS improvement by the combination strategy included nonsquamous cancer (P = 0.002), nonsmokers (P = 0.0005), and female (P = 0.02). | 2.0 | 0.173 |
| **N. Shokri-Mashhadi, 2023** | Concentration and clinical depressive disorder. | Melatonin supplementation | Placebo/other drugs | Bbrain-derived neurotrophic factor (BDNF) concentration and clinical depressive disorder. | Bbrain-derived neurotrophic factor (BDNF) concentration and clinical depressive disorder. | The subgroup analysis showed that melatonin supplementation had a significant decreasing effect on BDNF levels in doses ≤ 10 mg/day, with more than 4 weeks of duration, and in men. | 2.0 | 0.897 |
| **G. C. Siontis, 2016** | Severe aortic stenosis | Transcatheter aortic valve implantation | Surgival aortic valve replacement | All-cause mortality | All-cause mortalituy | In subgroup analyses, TAVI showed a robust survival benefit over SAVR for patients undergoing transfemoral access [0.80 (0.69-0.93); P = 0.004], but not transthoracic access [1.17 (0.88-1.56); P = 0.293] (P(interaction) = 0.024) and in female [0.68 (0.50-0.91); P = 0.010], but not male patients [0.99 (0.77-1.28); P = 0.952] (P(interaction) = 0.050) | 2.0 | 0.05 |
| **L. Sun, 2020** | Advanced metastasis | Anti-PD-1/PD-L1 inhibitors | Other drugs/placebo | Overall survival and progression free survival | Overall survival and progression free survival | Anti-PD-1/PD-L1 inhibitors were identified as a preferable treatment option for advanced or metastatic cancer patients who are male. | 2.0 | 0.88 |
| **M. J. Tarrahi, 2021** | Healthy subjects | Chromium supplementation | Placebo | Blood lipids | 1. Triglyceride Levels 2. HDL-C 3. LDL-C | older and non-obese subjects (>54 years and ≤ 29 kg/m2, respectively), women, Asian and Australian and picolinate form for TG, short-term, low dose, non-obese subjects, women, and Asian for VLDL, and nicotinate form for HDL-C, but had no effect on LDL-C. | 2.0 | TG- 0.64 LDL-C-0.42 HDL-C--0.17 |
| **L. Wu, 2017** | Blood pressure | Calcium + Vitamine D | No intervention/energy restriction | Blood pressure | Systolic blood pressure | Subgroup analysis by gender indicated some evidence of elevated SBP in male participants, and the WMD (95% CI) was 1.49 mm Hg (1.03, 1.95). | 2.0 | 0.1 |
| **K. Zhou, 2020** | Non-small cell lung cancer | Erlotinib + bevacizumab | Monotherapy | overall survival (OS), progression-free survival (PFS), objective response rate (ORR), and adverse events. | PFS | This improvement was especially notable in patients with the following characteristics: Eastern Cooperative Oncology Group Performance Status score of 0 or 1, female, | 2.0 | 0.7 |
| **S. K. Myung, 2021** | Cardiovascular disease | Calcium Supplements | Placebo | Cardiovascular disease (CVD), coronary heart disease (CHD) and cerebrovascular disease | CVD and CHD | The current meta-analysis found that calcium supplements increased a risk of CVD by about 15% in healthy postmenopausal women. | 2.0 | CVD-0.2 CHD-0.2 |
| **M. Vajdi, 2023** | Inflamation | Alpha-lipoic acid | Placebo/control | changes in CRP, IL-6, and TNF-α levels) | CRP, IL-6, | In subgroup analysis, ALA dosage, baseline concentrations of the parameter, sample size, and gender were considered as possible sources of heterogeneity. | 2.0 | CRP-0.384 IL-6-0.28 |
| **X. Niu, 2018** | Cryptogenic stroke | Transcatheter device closure (TDC) plus anti-thrombotic drugs | Medical management | Composite endpoint of ischemic stroke or transient ischaemic attack | ischemic stroke or transient ischaemic attack | The results indicated that participants with male gender, of younger age and with substantial shunt size significantly benefit from closure operation as compared to those with female gende | 2.0 | 0.19 |
| **S. Rastgoo, 2023** | Healthy or unhealthy adults | Conjugated linoleic acid | Placebo/control diet | Changes in CRP, IL-6, TNF, adiponectin, and leptin) | Adiponectin | Conjugate Linolenic acid decreases levels of adiponectin and leptin in women | 3.0 | 0.59 |
| **A. Granero-Gallegos, 2020** | Athletes | HRV. Controlled indurance training | Other forms of training | VO2max | VO2max | According to the sex subgroups (‘men vs. women’ vs. ‘men and women’), there were statistically significant improvements (p < 0.0001) in the three subgroups and statistically significant differences (p < 0.0001) between the three subgroups in favor of the women (men, ES = 0.33; women, ES = 0.40; men and women, ES = 0.19) | 3.0 | 0.53 |
| **L. Fan, 2023** | Gastric carcinoma | Programmed death 1 inhibitors plus chemotherapy | Chemotherapy | Overall survival and progression free survival | Overall survival | In this study we found that male patients (HR=0.78; 95% CI: 0.73–0.83, P<0.01) had a greater OS benefit with PD-1 inhibitors plus chemotherapy than female patients (HR=0.81; 95% CI: 0.74–0.89, P<0.01). A | 3.0 | 0.51 |
| **Zhan and Ho, 2005** | Healthy subjects | Soy protein containing isoflavones | Control diet | Changes in serum lipid concentrations | Changes in serum lipid concentrations | Soy protein containing isoflavones has a significant effect on serum lipid concentrations | 3.0 | 0.39 |
| **T. Tan, 2022** | Healthy subjects | Tai chi | Sedentary life | Cardiorespiratory fitnes outcomes | O2 pulse | The subgroup analysis suggested that overall in those who practiced Tai Chi, males (WMD = 1.48, 95% CI: 0.85 to 2.12, ) had higher O2 pulse than females (WMD = 0.73, 95% CI: 0.33 to 1.12, ). | 3.0 | / |
| **R. M. Hoffman, 2020** | Lung Cancer | Low-dose CT screening | Other screening techniques | Lung Cancer outcomes | Lung Cancer Death | LDCT screening significantly reduced LC mortality, though not overall mortality, with women appearing to benefit more than men. | 3.0 | 0.11 |
| **G. Jin, 2024** | Condyloma acuminatum | Paiteling | Other treatment | Clinical effectiveness | Clinical effectiveness | Subgroup analysis showed that the type of physical therapies, gender, and wart location might be the primary sources of heterogeneity. | 3.0 | 0.84 |
| **C. W. Liang, 2023** | Obesity | Linoleic acid | Usual Care | Body Composition | Body Composition | In subgroup analysis, the following factors were associated with significant outcomes: (1) body mass index ≥25 kg/m2; (2) female sex; | 5.0 | 0.09 |
| **Xu et al., 2022** | Disorders of consicousness | transcranial direct current stimulation | Placebo | Behavioral improvements | Behavioral improvements | he subgroup analysis showed that patients who were male or with a minimally conscious state (MCS) diagnosis were associated with a greater improvement in CRS-R score. | 5.0 | 0.002 but we recalculated and was 0.12 |
| **N. Amiri, 2021** | Healthy subjects | Resistance training (RT) | No trainning | muscle strength, insulin-like growth factor-1, and insulin-like growth factor-binding protein-3 | IGF-1 | Subgroup analysis revealed that the elevation in serum IGF-1 levels after RT was significant only in women (MD: 19.30 ng/ml); | 5.0 | 0.57 |

### Table S5. General information of claims without the ability to test for presence or not of statistical support

| **Author, year** | **Condition** | **Intervention** | **Control** | **Outcome of interest** | **Fallacious claims** | **Abstract Claim** | **Fallacy category** | **P-value** |
| --- | --- | --- | --- | --- | --- | --- | --- | --- |
| **S. Peng, 2022** | Healthy college students | E-health intervention | Other or no intervention | Change in physical activity (PA) and excessive sedentary behavior | Physical Activity | At follow-up, interventions in groups of developing region (SMD = 1.17, 95% CI: 0.73, 1.62, p < 0.001), objective instrument (SMD = 0.83, 95% CI: 0.23, 1.42, p = 0.007), duration ≤ 3-month (SMD = 1.06, 95% CI: 0.72, 1.39, p < 0.001), and all female (SMD = 0.79, 95% CI: 0.02, 1.56, p = 0.044) can significantly improve PA. | 1.0 | / |
| **S. Peng, 2021** | Atrial Fibrilation | Dexmedetomidine | Other/placebo | Atrial Fibrilation | Postoperative atrial fibrilation | Subgroup analysis showed that Dex was associated with reduced risk of atrial fibrillation after cardiac surgery in studies with smaller proportion of males (≤ 74%, OR = 0.55, 95% CI: 0.36–0.83, p = 0.005), but not in studies with older patients or larger proportion of males (p for subgroup difference = 0.02 and 0.04). | 1.0 | / |
| **H. Pei, 2014** | Peroperative Cardiorenal Events | Remote ischemic preconditioning | Placebo/non-placebo | Perioperative Cardiac and Renal Events | Perioperative Cardiac and Renal Events | A subgroup with more than 75% of male subjects (OR = 0.54; 95% CI, 0.38 to 0.76) has a more profound effect size than that with less than 75% of male ones (OR = 0.90; 95% CI, 0.68 to 1.20)(P = 0.02 for subgroup difference; Table 3). | 1.0 | / |
| **L. E. Miller, 2017** | Symptomatic tendinopathy | Platelet-rich plasma injection | Saline or anaestetic and or corticosteroids | Tendinopathy pain severity | Tendinopathy pain severity | In subgroup analysis and meta-regression, studies with a higher proportion of female patients were associated with greater treatment benefits with PRP. | 1.0 | / |
| **L. E. Miller, 2013** | Healthy or constipated adults | Probiotic supplementation | Control | Intestinal transit time | Intestinal transit time | Constipation (r (2) = 39%, P = 0.01), higher mean age (r (2) = 27%, P = 0.03), and higher percentage of female subjects (r (2) = 23%, P < 0.05) were predictive of decreased ITT with probiotics in meta-regression. | 1.0 | / |
| **F. Meng, 2018** | Brucellosis | Rifampicin | Streptomycin | Brucellosis | Brucellosis | Men with brucellosis who received rifampicin had a higher failure risk than women | 1.0 | / |
| **S. Matsunaga, 2019** | Alzheimers | Idalopirdine | Placebo | Alzheimer's Disease Assessment Scale-cognitive subscale | Alzheimer's Disease Assessment Scale-cognitive subscale | Regarding the meta-regression analysis of the proportion of male participants, drug effects may have been more significant in the group with a high proportion of female participants. In our meta-analysis, studies with higher proportion of female participants tend to have higher baseline ADAS-cog scores than studies with lower proportion of female participants. Therefore, the results of this meta-regression analysis may be related to the severity of dementia | 1.0 | / |
| **M. Magill, 2009** | Alcohol/drugs addiction | Cognitive-behavioral treatment | No treatment | Substance use | Substance use | Meta-regression analyses indicated that the percentage of female participants was positively associated and the number of treatment sessions was negatively associated with effect size. | 1.0 | / |
| **C. Lin, 2020** | Diabetes Mellitus | Drug | Placebo | Placebo response | Placebo response | However, younger age (β = 0.02, 95% CI, 0.01 to 0.03, P = 0.01), lower male percentage (β = 0.01, 95% CI, 0.22 × 10(-2), 0.01, P < 0.01), higher baseline BMI (β = - 0.02, 95% CI, - 0.04 to - 0.26 × 10(-2), P = 0.02), and higher baseline HbA1c (β = - 0.09, 95% CI, - 0.16 to - 0.01, P = 0.02) were significantly associated with more HbA1c reduction by placebo in T2DM | 1.0 | / |
| **X. Li, P. Chang, 2020** | Cardiovascular disease | ACE inhibitors | Other antihypertensives | Arterial stifness | Arterial stifness | subgroup analyses indicated that the ACEIs were associated with increased levels of ba-PWV if the study was conducted in Western countries (WMD: 1.02; 95% CI: 0.17 to 1.87; P=0.019), mean age of <60.0 years (WMD: 0.71; 95% CI: 0.04 to 1.39; P=0.037), percentage male ≥60.0% (WMD: 1.02; 95% CI: 0.17 to 1.87; P=0.019), compared with ARB (WMD: 0.71; 95% CI: 0.04 to 1.39; P=0.037), | 1.0 | / |
| **M. S. Kim, 2017** | Postoperative Nausea and Vomiting | Palonosetron | Ramosetron | Postoperative Nausea and Vomiting | Postoperative Nausea and Vomiting | However, palonosetron was more effective in preventing POV, for females and laparoscopies (RR, 0.56; 95% CI, 0.36 to 0.86; p=0.009 and RR, 0.46; 95% CI, 0.23 to 0.94; p=0.033). | 1.0 | / |
| **T. Hong, 2019** | Knee pain | Invasive Radiofrequency Treatment | Control | Knee pain and function | Knee pain and function | The meta-regression analysis further proved that the higher the female proportion, the better the treatment effect, although the underlying mechanism is yet to be elucidated. Women are prone to suffering from chronic pain syndrome, and the frequency, areas of the body, duration, and severity of pain are significantly higher than those in men [44–46] | 1.0 | / |
| **Y. Fang, 2012** | Bone density | Vitamin K | Other suplements/glucocorticoids or placebo | Bone density | Bone density | Subgroup analysis revealed that ethnic difference, gender, and vitamin K type were associated with variable effects on BMD at the lumbar spine. | 1.0 | / |
| **X. Chen, 2017** | Fibromyalgia | Control placebo | Untreated | Placebo response | Placebo response | respect to gender, the more women included in a trial, the less effective was placebo. Trials with <80% women had a placebo ES of 0.65 (95%CI 0.32 to 0.98), whereas trials with 100% women had a placebo ES of 0.21 (95%CI 0.02 to 0.39). | 1.0 | / |
| **C. H. Chang, 2022** | Methamphetamine addiction | Repetitive Transcranial Magnetic Stimulation | Sham | Craving Methamphetamine | Craving Methamphetamine | A metaregression revealed that the SMDs increased with the increase in baseline craving scores, whereas they decreased with the increase in the proportion of men and duration of abstinence. | 1.0 | / |
| **C. H. Chang, 2019** | Schizophrenia | NMDA-receptor-enhancing agents | Placebo | Cognitive outcomes | Cognitive outcomes | Men were associated with a smaller effect of NMDA-receptor-positive modulators on overall cognitive function. | 1.0 | / |
| **E. K. Calton, 2017** | Systemic inflamation | cholecalciferol supplementation | Placebo | Systemic inflammatory profile | Systemic inflammatory profile | Age (β=−0.06, 95% CI −0.103 to −0.017, P=0.01), gender (β=0.027, 95% CI 0.011–0.044, P=0.004) and supplementation duration (β=0.049, 95% CI=0.018–0.079, P=0.005) were significant predictors of CRP change (Table 3). | 1.0 | / |
| **M. Cai, 2016** | Obesity | 1. Aerobic Exercise 2. Aerobic Exercise + Diet | 1. No intervention 2. Diet | Lipids and glucose profile | Lipids and glucose profile | Subgroup analyses showed that AE significantly changed the HDL-C level in female population and when intervention protocol of AE+diet vs. diet only was used. | 1.0 | / |
| **J. R. Beguerie, 2010** | Melanoma | Tamoxifen | Non-Tamoxifen | Response and mortality | Response and mortality | Subgroup analyses showed that female patients were more likely to respond. | 1.0 | / |
| **H. Bakouni, 2023** | Amphetamine addiction | Bupropion | Placebo | Amphetamine-type stimulant craving | Amphetamine-type stimulant craving | greater reduction in end-of-treatment ATS craving in studies with mixed ATS use frequency (SMD: -0.46; 95%CI: -0.70, -0.22) and male-only samples (SMD: -1.26; 95%CI: -1.87, -0.65). | 1.0 | / |
| **H. Huang, 2021** | Perioperative anxiety | Aromatherapy | Standard care/placebo | Self rated anxiety | Perioperative anxiety | most subg/roups showed a significant effect of aromatherapy on preoperative anxiety, except for the no treatment subgroup (MD: 5.40, 95%CI: 7.76 to 0.71) and female subgroup (MD: 3.96, 95%CI: 9.19 to 1.27). CONCLUSION: | 1.0 | / |
| **A. T. Olagunju, 2018** | Schizophrenia | Clozapine | Other antipsychotics | Psychosocial function | Psychosocial function | Baseline severity of illness, illicit drug use, extrapyramidal side effects, sex and cognition explained the variability in functional outcome | 1.0 | / |
| **H. E. Brown, 2013** | Depression | Physical Activity Interventions | Non-physical control or control | Depression | Depression | Subgroup analyses showed that methodological (e.g. studies with both education and PA intervention; those with a higher quality score; and less than 3 months in duration) and participant characteristics (e.g. single-gender studies; those targeting overweight or obese groups) contributed most to the reduction in depression. | 1.0 | / |
| **L. E. Miller, 2016** | Adult population | Probiotic supplementation | Non-probiotic supplementation | Intestinal transit time | Intestinal transit time | Constipation (R (2) = 38%, P < 0.01), higher study quality (R (2) = 31%, P = 0.01), older age (R (2) = 27%, P = 0.02), higher percentage of female subjects (R (2) = 26%, P = 0.02), | 1.0 | / |
| **Yuan et al., 2022** | Gastric carcinoma | Immune checkpoint inhibitors (ICIs) | Chemotherapy, Placebo | Treatment-Related Adverse Events (TRAEs) | Treatment-Related Adverse Events (TRAEs) | Significant correlation between all grade TRAEs and proportion of female patients | 1.0 | / |
| **Zhang et al., 2021** | Frozen Shoulder | Nonsurgical interventions for frozen shoulder | Placebo | Pain relief, shoulder function improvement, range of motion | Pain relief, shoulder function improvement, range of motion | Female sex and diabetes identified as moderators of effectiveness | 1.0 | / |
| **Zhang et al., 2022** | Headache | Pharmacologic treatments for primary headaches | Placebo | Nocebo responses in primary headache treatments | Nocebo responses in primary headache treatments | Nocebo responses were influenced by several factors including female sex | 1.0 | / |
| **Zhao et al., 2017** | Hypothyroidism | Levothyroxine (L-T4) therapy | Placebo | Carotid intima-media thickness (C-IMT) | Carotid intima-media thickness (C-IMT) | L-T4 therapy decreases C-IMT in patients with subclinical hypothyroidism (SCH), more pronounced in female patients | 1.0 | / |
| **Amiri et al., 2022** | Obesity | Motivational interviewing (MI) for weight management | Various other interventions or no intervention | BMI, BMI Z-score, waist circumference, fat percentage | Central obesity | A reduction in central obesity was noted predominantly among females | 1.0 | / |
| **Borek, 2018** | Obesity | Group-based weight loss interventions | Not clear | Weight loss | Weight loss | Explicitly targeting weight loss, men-only groups providing feedback and dietary goals were significantly associated with greater effectiveness (p < .05). | 1.0 | / |
| **J. Schneider-Thoma, 2018** | Mental Disorders | Second-generation antipsychotic drugs | Placebo | Short-term mortality | Short term mortality | The exceptions were increased mortality in patients with dementia (OR 1·56; 95% CI 1·10-2·21), in elderly patients (1·38; 1·01-1·89), in aripiprazole-treated patients (2·20; 1·00-4·86), and in studies with a higher proportion of women (regression coefficient 0·025; 95% credible interval 0·010-0·040). | 1.0 | / |
| **J. W. Senefeld, 2020** | Healthy subjects | Nitrate supplementation | Placebo | Exercise performance | Exercise performance | Subgroup analyses conducted on biological sex, aerobic fitness, and FiO2 demonstrated that the ergogenic effect of NO3 supplementation was as follows: 1) not observed in studies with only women (n = 6; d = 0.116; 95% CI, -0.126 to 0.358; P = 0.347), 2) n | 1.0 | / |
| **M. Sun, 2023** | Severe COVID-19 | Molnupiravir | Multiple outcomes | Elevated risk of hospitalization | Viral clearance & risk of hospitalization | For the rate of viral clearance, subgroup effects were found between trials with low and high risk of bias (P = 0.001) and between trials with male or female majority (P < 0.001). For admission to hospital, subgroup effects were also found between trials with ≥50% and <50% of the participants being female (P = 0.04). Meta-regression showed a significant association between higher trial mean age and elevated risk of hospitalization (P = 0.011), and female majority and elevated risk of hospitalization (P = 0.011). | 1.0 | / |
| **J. K. Tshiananga, 2012** | Diabetes Mellitus | Nurse-led Diabetes Self-management Education (DSME) | Usual Care (UC) | Glycosylated Hemoglobin (A1C), Blood pressure, Lipid levels | Glycaemic control and cardiovascular risk factors | Nurse-led DSME improves glycemic control and cardiovascular risk factors, with significant effects among males. | 1.0 | / |
| **C. Zhou, 2012** | Primary coronary intervention | Ischaemic postconditioning | Control | Myocardial enzyme levels and LVEF | Myocardial enzyme levels and LVEF | Availableevidencefromthissystematicreviewandmeta-analysisof10RCTssuggeststhat IPoCmayconfercardioprotection intermsofmyocardial enzyme levelsandLVEF forSTEMIduringprimaryPCI.Theseeffectsaremore pronouncedamong young andmalepatients, and those inwhomdirect-stenting techniqueswereused | 1.0 | / |
| **H. Azizi, 2022** | Suicide | Brief Contact Interventions | Treatment as usual | Re-attempt | Meta-regression analysis explored that BCI time (more than 12 months), BCI type, age, and female sex were the potential sources of the heterogeneity. | Meta-regression analysis explored that BCI time (more than 12 months), BCI type, age, and female sex were the potential sources of the heterogeneity. | 1.0 | / |
| **Sh. Bai, 2020** | Major depressiove disorder | anti-inflamatory agents | Placebo | Depresive symptoms | depressive symptoms | For women-only trials, no difference in changes of depression severity was found between groups | 1.0 | / |
| **Z. A. Bhutta, 2000** | Persistent Diarrhea | Oral zinc | Control | Persistent diarrhea | Persistent diarrhea | In none of the subgroup analyses were the 2 subgroups of each pair significantly different from each other; however, in persistent diarrhea there tended to be a greater effect in subjects aged <12 mo, who were male, or who had wasting or lower baseline plasma zinc concentrations. | 1.0 | / |
| **Yun et al., 2017** | HIV | PrEP (pre-exposure prophylaxis) | Placebo | Medicine-taking compliance (MTC) and HIV incidence | Medicine-taking compliance (MTC) and HIV incidence | Female and younger subjects have lower adherence in PrEP trials | 1.0 | / |
| **J. Grgic, 2022** | - | Caffeine | NO info | Isometric handgrip strength | Isometric handgrip strength | these ergogenic effects were very small and were observed mostly among male participants. | 5.0 | / |
| **J. Fernández-Guisasola, 2010** | Postoperative Nausea and Vomiting | Nitrous oxide | No nitrous oxide | Postoperative nausea and vomiting in adults | Postoperative nausea and vomiting in adults | The maximal risk reduction was obtained in female patients (pooled relative risk0.76, 95% CI 0.60–0.96) | 5.0 | / |
| **C. H. Wang, 2012** | UTI | Cranberry containing products | Placebo or no placebo | Incidence of urinary tract infections (UTI) | Incidence of UTI | On subgroup analysis, cranberry-containing products seemed to be more effective in several subgroups, including women with recurrent UTIs (RR, 0.53; 95% CI, 0.33-0.83) (I(2) = 0%), female populations (RR, 0.49; 95% CI, 0.34-0.73) (I(2) = 34%), | 5.0 | / |
| **C. Wang, 2015** | Osteoporosis | Paratyhroid Hormone + Alendronate | Other treatment | Mean percent increase in bone mass density of lumbar spine, femoral neck, total hip, and distal readius | Spine | Subgroup analysis revealed that among the patients in the combination therapy group, greater increases in the spine BMD were observed when the PTH was administered with a dosage of 20 μg (WMD = 2.33, 95%CI: 1.24, 3.43; p = .000), or the treatment duration lasted more than 12 months (WMD = 2.23, 95%CI: 1.00, 3.47; p = .000), or the combination therapy was used in osteoporosis women (WMD = 1.58, 95%CI: 0.63, 2.53; p = .001). | 5.0 | / |
| **P. Sarraf, 2019** | Healthy adults | Curcumin | Placebo | Brain-derived neutrophic factor | Brain-derived neutrophic factor | Subgroup analysis showed that sex, mean age of participants, curcumin dosage, and trial duration were potential sources of heterogeneity. | 5.0 | / |
